# Deep Learning for Freezing of Gait Assessment using Inertial Measurement Units: A Multicentre Study

**DOI:** 10.1101/2025.06.27.25330405

**Authors:** Po-Kai Yang, Juha Carlon, Maaike Goris, Emilie Klaver, Jorik Nonnekes, Richard J. A. van Wezel, Lisa Alcock, Alison J. Yarnall, Lynn Rochester, Clint Hansen, Christian Schlenstedt, Walter Maetzler, David Buzaglo, Marina Brozgol, Jeffrey M. Hausdorff, Alice Nieuwboer, Moran Gilat, Pieter Ginis, Bart Vanrumste, Benjamin Filtjens

## Abstract

Video annotation is the gold-standard method to assess Freezing of Gait (FOG) in Parkinsonian disorders, but it is time-consuming. Deep learning (DL)-based assessment of FOG using inertial measurement units ameliorates these problems but poses challenges. Particularly, the large heterogeneity between patients and assessment methods potentially affects detection performance between independent cohorts. To evaluate heterogeneity effects, we developed a DL model on a local cohort (85 participants; 2043 trials) and validated it across six external cohorts (256 participants; 1058 trials). Model-expert agreement on the percentage-of-time-frozen was strong locally (ICC=0.886 [0.79,0.90]) but reduced in external cohorts (ICC=0.562***±***0.141). Fine-tuning the DL model with just 50 minutes of external cohort data improved the ICC to 0.732***±***0.138, falling within the borderline of the inter-rater agreement (ICC=0.73-0.99). Therefore, while unified standards are still being developed, we propose an expert-in-the-loop workflow as an effective intermediary and present a proof-of-concept web-based platform for fine-tuning and expert review (aidfog.be).

## 1 Introduction

Freezing of gait (FOG) is a debilitating motor symptom observed predominantly in people with Parkinson’s disease (PD) and related disorders Kihlstedt et al (2024); Factor (2008). FOG is currently clinically defined as a “brief, episodic absence or marked reduction of forward progression of the feet despite the intention to walk and reach a destination” Nutt et al (2011). FOG significantly compromises mobility, independence, and quality of life and increases the risk of falls Moore et al (2005); Bloem et al (2004); Giladi and Hausdorff (2006). It is estimated that FOG affects up to 80% of people over the course of PD Perez-Lloret et al (2014); Hely et al (2008), highlighting its substantial prevalence and clinical relevance.

The gold standard to assess the presence of FOG and severity involves dedicated gait tasks, so-called FOG-provoking tasks, that challenge dynamic motor-cognitive or motor-sensory control by including triggers that frequently elicit FOG (e.g., gait initiation, narrow passageways, cognitive dual tasks, and rapid 180° or 360° turning) Snijders et al (2008); D’Cruz et al (2021); Mancini et al (2019); Zoetewei et al (2024); Goris et al (2025). Camera recordings capture these gait tasks, which are then meticulously reviewed by trained evaluators to annotate individual FOG episodes (Figure 1a) Gilat (2019). This method allows FOG severity to be quantified as the percentage of time spent with FOG compared to the total task duration (%TF) Morris et al (2012), and the number of FOG episodes (#FOG). However, this process is extremely labor-intensive, requiring trained evaluators to manually review video footage on a frame-by frame basis, which slows the overall assessment pace. Additionally, human judgment introduces variability across evaluators Cockx et al (2022), which complicates the standardization of FOG assessment and impacts the reliability of determining the treatment response Lewis et al (2022); Barthel et al (2016); Mancini et al (2019).

**Fig. 1:**
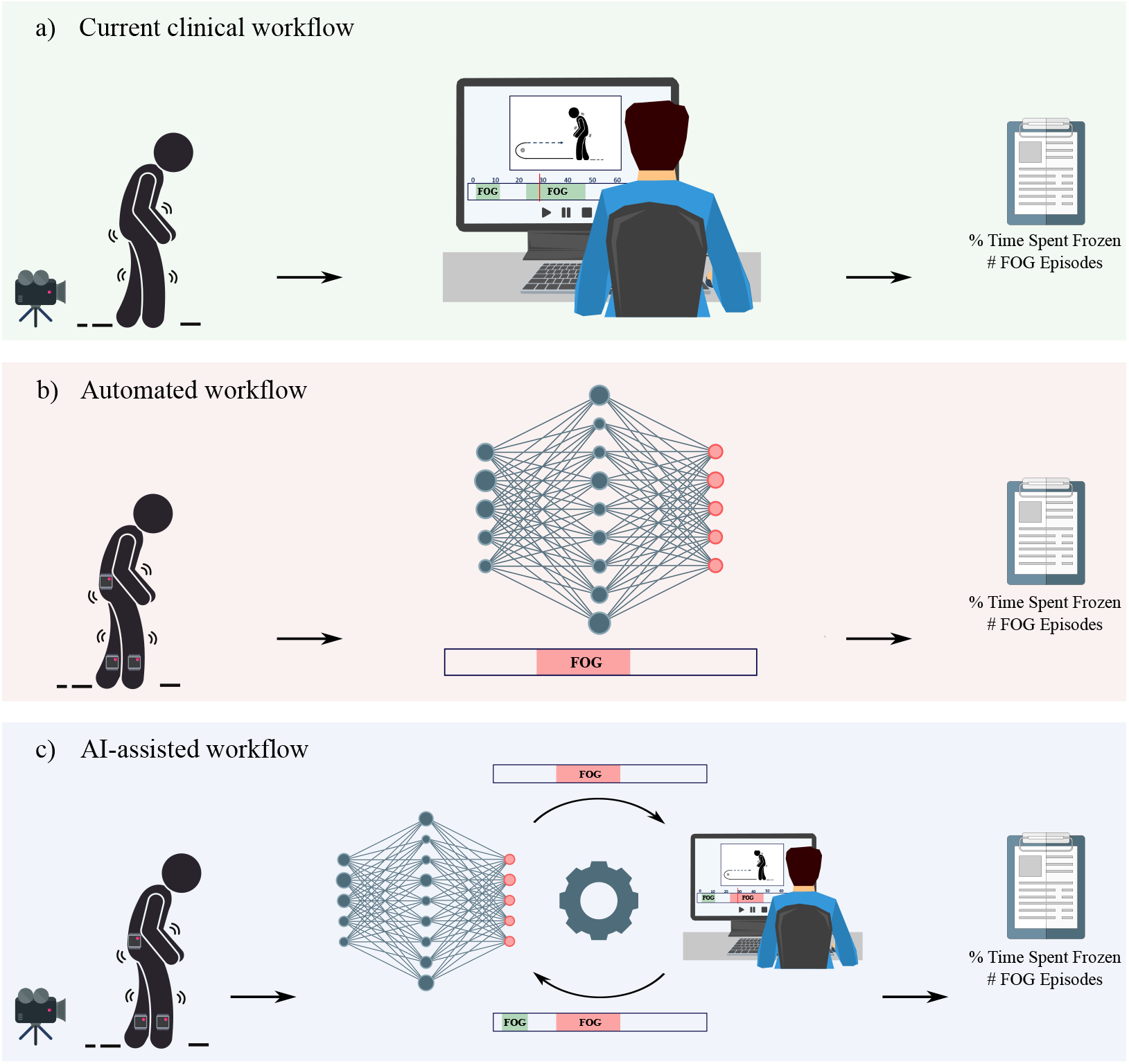
In the current clinical workflow (a), trained human evaluators review camera footage of gait tasks to manually identify freezing of gait (FOG) episodes. In an automated workflow (b), gait data collected from inertial measurement units (IMUs) are processed by a deep-learning algorithm, which automatically identifies potential FOG episodes. In an AI-assisted “expert-in-the-loop” workflow (c), the deep-learning algorithm provides an initial evaluation of the gait task using IMU data. A human evaluator can then review the annotations and refine them as needed, incorporating additional information from camera footage. The refined predictions can subsequently be used to fine-tune the deep-learning algorithm. In all workflows, FOG severity is quantified using outcome measures derived from the identified FOG episodes.

Recent advances in machine learning, particularly driven by deep learning (DL), combined with the ubiquity and affordability of wearable sensors such as inertial measurement units (IMUs), hold promise to overcome the limitations of manual FOG annotation (Figure 1b). Prior research has highlighted that DL algorithms (DLA) when combined with IMUs, enable reliable assessment of FOG Mancini et al (2019, 2025b). Nevertheless, the reliability of these automated analyses was constrained by the possible pitfall of overfitting the model to detect such a complex and heterogeneous symptom when only limited sample sizes are available or diminished reproducibility stemming from the absence of multicentre datasets Qi et al (2025); Mancini et al (2025b). Consequently, DLAs trained on a specific dataset may not generalize well to detect FOG in other datasets. In this field, poor generalization between datasets is driven by four key sources of variability: (1) task differences, where FOG-provoking tasks involve movements or types of volitional stopping that differ from those in training data; (2) annotation criteria, as the current FOG definition leaves room for subjective interpretation, making its consistent operationalization for annotation challenging; (3) variations in IMU configurations, including differences in sensor numbers and placements; and (4) patient spectrum bias, as most FOG detection studies focus on severe freezers, making generalization to mild freezers uncertain. These inconsistency issues prompted the International Consortium for FOG (ICFOG)^1^ to work towards standardized measurement protocols Mancini et al (2025b); Lewis et al (2022); Salomon et al (2024). Moreover, integration of DLAs into a clinical workflow could face obstacles due to the limited interpretability of the models, often referred to as the “black box” phenomenon Wang et al (2020). One possible solution is a human-in-the-loop approach, in which clinicians review the model’s predictions and the rationale behind them (e.g., by determining which data patterns influenced a decision) before using those predictions to guide treatment, thereby building trust and transparency Mosqueira-Rey et al (2023); Kumar et al (2024).

To address these challenges and build on our earlier IMU-based FOG detection study using a Multi-Stage Temporal Convolutional Network (MS-TCN) Yang et al (2024), we proceeded in three steps. First, we evaluated three state-of-the-art sequence learning architectures, including MS-TCN Farha and Gall (2019), the Action Segmentation Transformer (ASFormer) Yi et al (2021a), and the Long-Term Context Action Segmentation model (LTContext) Bahrami et al (2023), to identify the best model for FOG severity assessment across external datasets and quantify how cohort differences affect model performance. Next, to address possible cohort differences, we compared an adaptation strategy on the external datasets, i.e., by fine-tuning the model with varying amounts of annotated data, and comparing this with retraining a model from scratch. Finally, to encompass model fine-tuning and support an “expert-in-the-loop” approach to potentially increase trust in the model prediction, we introduced an AI-assisted workflow (Figure 1c), where the DLA supports but does not replace researchers in annotating FOG episodes. We envision this workflow to be supported by an interactive proof-of-concept web-based platform that could serve two purposes: (1) enhancing transparency and trust by enabling researchers to review and refine model predictions; and (2) fine-tuning the DLA using these refined predictions, thereby improving its generalization to data collected in different centers. To operationalize this workflow and facilitate future user studies, we have built the AID-FOG (AI-Driven Freezing of Gait) platform, a proof-of-concept web application that integrates expert review and iterative model fine-tuning. For a detailed design process of the platform and a live demo, refer to Supplementary Appendix H.

## 2 Results

### 2.1 Dataset and participant characteristics

We included datasets from seven cohorts collected across eight universities. The local cohort comprised four datasets from various KU Leuven projects, including those used in previous studies Yang et al (2024); Spildooren et al (2010); Vervoort et al (2016), as well as data from an ongoing study (ClinicalTrials.gov NCT05511597). These were collectively used as the local training dataset. The six external cohorts included: (1) a non-public dataset from Radboud University Janssen et al (2017), (2) a public dataset from Stanford University O’Day et al (2022), (3) a public dataset from the Federal University of ABC Ribeiro De Souza et al (2022), (4) a public dataset from the University of Twente Delgado-Terán (2025); Delgado-Terán et al (2025), (5) a public dataset from Beijing Xuanwu Hospital Zhang et al (2022); Li (2021), and (6) a non-public multicenter dataset from the Mobilise-D project Goris et al (2025), which includes data from KU Leuven, University Medical Center Schleswig-Holstein Campus Kiel, and Tel Aviv Sourasky Medical Center. As potential sources of variability, the Stanford and Radboud cohorts employed different FOG-provoking tasks; the Federal, Twente, and Xuanwu cohorts introduced variations in the number of IMU sensors; and the Mobilise-D cohort reflected differences in patient spectrum. The FOG annotation criteria also varied slightly across cohorts and are detailed in Section 4.

Table 1 summarizes the patient and data characteristics across the seven cohorts. All participants had a clinical diagnosis of idiopathic PD. Participants varied in age and disease duration, with the Stanford cohort having the youngest average age (58 ± 5 years) and the Radboud cohort the oldest (72 ± 4 years). Among the external cohorts, Radboud and Stanford featured distinct FOG-provoking tasks, e.g., 15-meter walking with volitional stopping (Radboud) and slalom with figure-eight turns (Stanford), which differed from those used in the KU Leuven cohort, such as the TUG, 360° turning, 5-meter walk, and slalom tasks. Additionally, while most datasets used five IMUs, less IMUs was available for the Federal, Twente, and Xuanwu cohorts. As described in Section 4.2.1, as the original Stanford and Twente publications only specified “ankle-mounted” and “foot-mounted” sensors without naming the exact bones, we harmonized placements of ankle-mounted IMUs correspond to tibia positions and their foot-mounted IMUs to talus positions. Based on the New Freezing of Gait Questionnaire (NFOG-Q) and the Movement Disorders Society Unified Parkinson’s Disease Rating Scale (MDS-UPDRS) III scores, the Radboud, Federal, Twente, and Xuanwu cohorts exhibited similar disease and FOG severity to KU Leuven, while Mobilise-D had lower scores, indicating milder disease severity and lower probability for FOG Tan et al (2011). Indeed, the average FOG episode duration in Mobilise-D was the shortest among all datasets and only 0.983 seconds, suggesting a predominance of mild freezers experiencing very short FOG episodes.

**Table 1:**
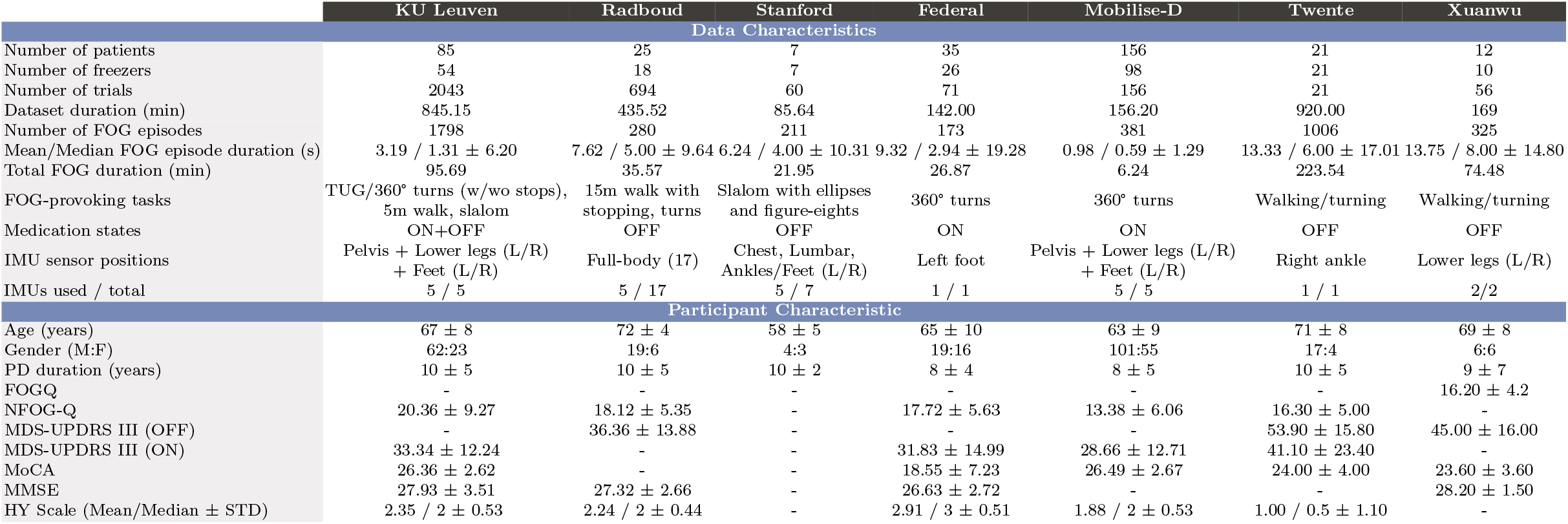
Dataset characteristics. FOG: Freezing of Gait, PD: Parkinson’s disease, FOGQ: Freezing of Gait Questionnaire, NFOG-Q: New Freezing of Gait Questionnaire, MDS-UPDRS: Movement Disorders Society Unified Parkinson’s Disease Rating Scale, MMSE: Mini-Mental State Examination, MoCA: Montreal Cognitive Assessment. *Note: MDS-UPDRS III scores were OFF only in the Radboud and Xuanwu dataset; in KU Leuven, Vervoort et al (2016) provided OFF scores and the remaining participants ON (we report only ON); the Federal and Mobilise-D cohorts provided ON only; and the Twente cohort provided both OFF and ON.

For model pre-training, 2043 gait trials from 85 participants were included in the KU Leuven cohort. The %TF was 11.32% (95.69 out of 845.15 minutes), with a total of 1798 FOG episodes observed. For external validation, 1058 trials from 256 participants across the six external cohorts were used. The %TF was 20.37% (388.65 out of 1908.36 minutes), with a total of 2376 FOG episodes recorded.

### 2.2 Pretrained model externally validated

We first compared three state-of-the-art sequence learning architectures for FOG detection, i.e., MS-TCN, ASFormer, and LTContext. As detailed in Supplementary Appendix C, LTContext outperformed the others on average and was selected for further analysis. In the local KU Leuven cohort, LTContext achieved strong agreement with experts on %TF (ICC = 0.886, 95 % confidence interval (CI) [0.79, 0.90]) and fair agreement on #FOG (ICC = 0.573, 95 % CI [0.28, 0.71]). In trials with at least one FOG episode, it achieved a sample F1 of 0.291 and a segment F1@50 of 0.123. In non-FOG trials, it made just 0.302 false positive (FP) episodes per trial and a mean FP duration per trial of 0.230 seconds, which helped maintain the ICC. These relatively low F1 scores, calculated at a strict, trial-wise level (see Section 4.4 for details), underscore the challenges of detecting FOG, even on local data. For a broader set of performance metrics (e.g., accuracy, recall, specificity, and precision) computed on the entirely dataset level, please refer to Supplementary Appendix G.

When tested on the external cohorts (Table 2), agreement in terms of FOG severity dropped: average ICC for %TF reduced to 0.562 ± 0.141 (95 % CI [−0.01, 0.78]) and for #FOG to 0.298 ± 0.270 (95 % CI [−0.16, 0.49]). The sample F1 in FOG trials increased slightly to 0.390 ± 0.162 (F1@50 = 0.185 ± 0.113), but the #FP in non-FOG trials increased to 1.690 ± 1.579 per trial (mean FP duration 5.076 ± 5.939 seconds). This results in a lowered agreement in both %TF and #FOG, showing that differences between cohorts can reduce model performance.

**Table 2:**
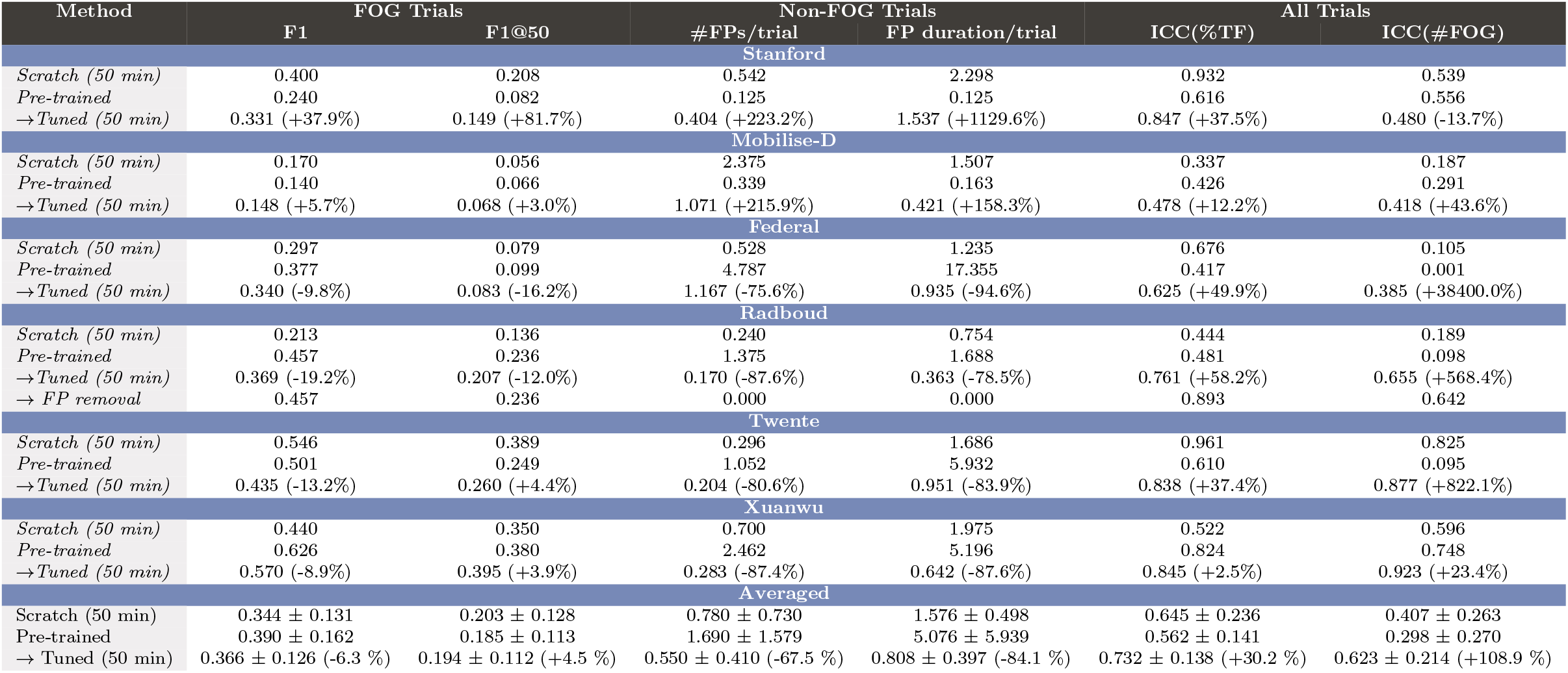
Performance of DLA models trained from scratch, pre-trained, and fine-tuned. Results are reported for each dataset using 50 minutes annotated data for fine-tuning and training-from-scratch. Overall performance is summarized as the arithmetic mean and standard deviation across datasets. Percentage improvements are calculated relative to the pre-trained model using 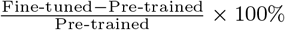:

### 2.3 Fine-tuned model externally validated

To address the performance drop observed in the external datasets, we explored fine-tuning as an adaptation strategy for the LTContext model pre-trained on the local KU Leuven cohort and compared it against training a new model with randomly initialized weights from scratch directly on the external datasets. As shown in Figure 2, fine-tuning the model with increasing amounts of data (from 10 to 360 minutes) consistently improved ICC scores for both %TF and #FOG compared to the pre-trained baseline model, with gains plateauing at around 50 min of training data. At that point, fine-tuned ICC(%TF) reached 0.732 ± 0.138 (+30.2 %), and ICC(#FOG) reached 0.623 ± 0.214 (+108.9 %). Models trained from scratch followed the same training trajectory when more annotated data was added but plateaued at ICC(%TF)=0.708± 0.195 after 90 min and ICC(#FOG)=0.528 ± 0.244 after 270 min. This result underscores that fine-tuning the pre-trained model is more data-efficient for adapting to external cohorts than training a new model from scratch.

**Fig. 2:**
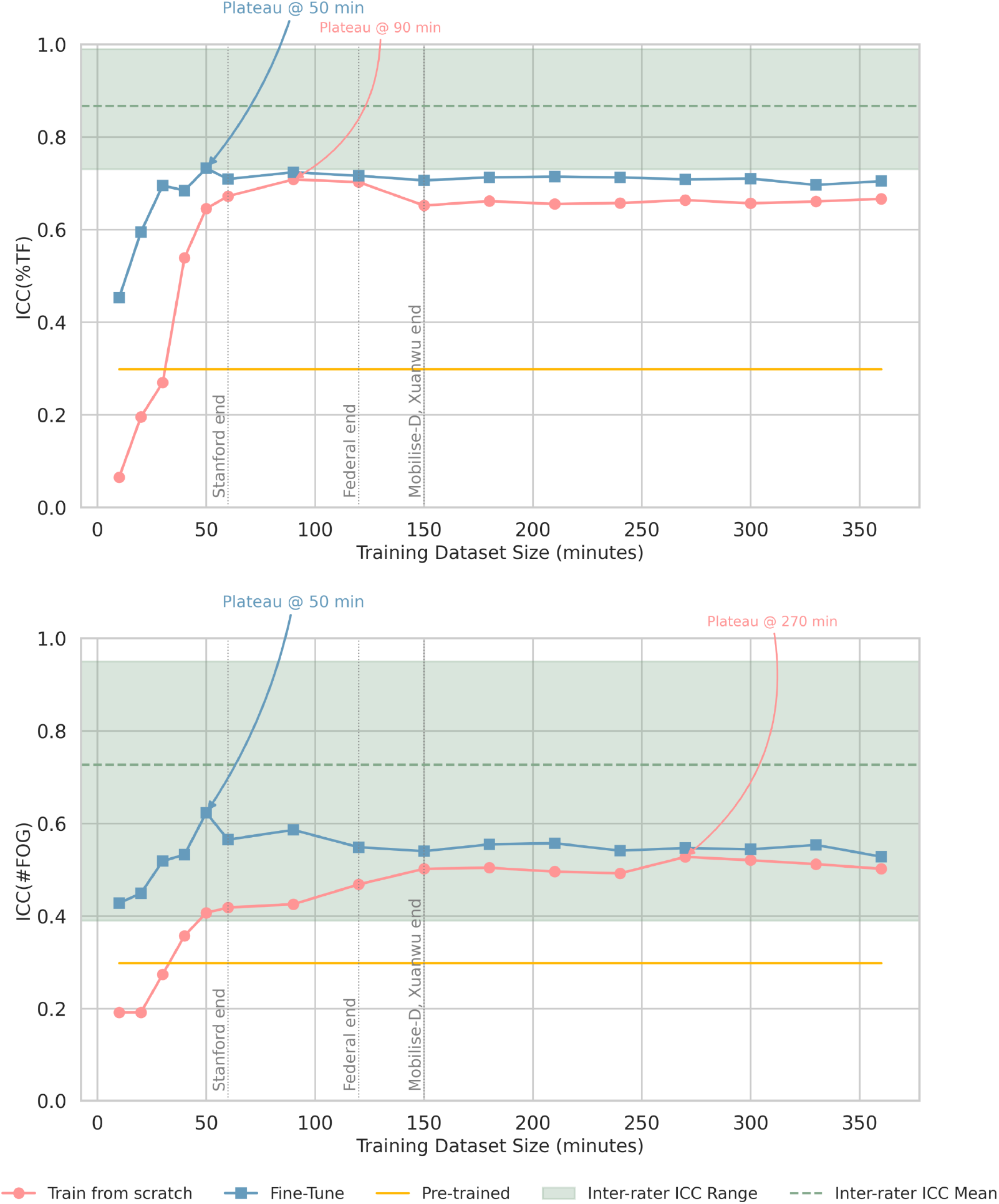
Average ICC(%TF) and ICC(#FOG) across six external cohorts for models trained from scratch (red), pre-trained (yellow), and fine-tuned (blue), plotted across varying training dataset size. Vertical gray lines indicate the time points beyond which data was no longer available for a given cohort. When a dataset lacked data at a specific time point, its last available value was used to compute the average, based on the common assumption that model performance generally improves, or at least does not deteriorate, with more training data Zhu et al (2016). The shaded green area represents the range (mean *±* min/max) of inter-rater ICCs for %TF and #FOG reported in previous studies O’Day et al (2022); Kondo et al (2022); Morris et al (2012); Mancini et al (2021); D’Cruz et al (2021). Notably, fine-tuning raises ICC(#FOG) into the inter-rater range (0.390-0.950), while ICC(%TF) remains slightly below (0.730-0.990), primarily due to lower improvement in the Mobilise-D cohort, which includes mostly mild freezers.

Nevertheless, as shown in Figure 2, even after fine-tuning or training from scratch, ICC(%TF) remained in the lower boundary of the ICC ranges between human raters reported in the literature (0.730-0.990 for %TF and 0.390-0.950 for #FOG). Among all cohorts, Mobilise-D showed the smallest gains, likely due to the difficulty of annotating shorter FOG episodes during 360° turning-in-place tasks in mild freezers (see cohort-difference analysis below).

Additionally, we identified four main challenges affecting generalization in FOG detection: (1) task differences (e.g., Radboud and Stanford), (2) annotation criteria (e.g., Stanford), (3) variations in IMU configurations (e.g., Federal, Twente, Xuanwu), and (4) patient spectrum bias differences (e.g., Mobilise-D). Our per-dataset analysis showed that fine-tuning mitigated cohort differences. On average, for challenge 1 (task differences), ICC(%TF) improved from 0.549 to 0.870 (+58.5%) and ICC(#FOG) from 0.327 to 0.568 (+73.7%). For challenge 2 (annotation criteria), ICC(%TF) increased from 0.616 to 0.847 (+37.5%), though ICC(#FOG) slightly declined from 0.556 to 0.480 (−13.7%). For challenge 3 (IMU variations), ICC(%TF) improved from 0.617 8 to 0.769 (+24.6%) and ICC(#FOG) from 0.281 to 0.728 (+159.1%). For challenge 4 (patient spectrum bias), ICC(%TF) increased from 0.426 to 0.478 (+12.2%) and ICC(#FOG) from 0.291 to 0.418 (+43.6%). Here, we report ICCs to support clinical interpretation. In Supplementary Appendix F, we demonstrate how fine-tuning on each dataset changes sample- and segment-level F1, mean #FP per trial, and mean FP duration per trial, and how these metric changes drive the observed ICC values.

Figure 3a shows a Stanford example where experts annotated shuffling as FOG. These subtle IMU patterns were not initially detected by the pre-trained model but were correctly identified after fine-tuning, though this also led to an overall increase in FPs (false positives). Figure 3b shows an example from Radboud, which includes short pre-task standing phases (e.g., before 2 seconds) and intermediate stopping periods (e.g., 60-65 seconds). The pre-trained model misclassified these as FOG, but fine-tuning enhanced its ability to distinguish volitional stops from true FOG episodes. Similarly, Figures 3c, 3d, and 3f illustrate the model’s tendency to generate multiple FPs in datasets with only one or two IMUs, i.e., Federal, Twente, and Xuanwu, potentially due to zero-filling of missing channels to match the 5-IMU input of the pretrained model. Fine-tuning reduced these FPs, with further improvements observed as more annotated data were used for training. Figure 3e illustrates the challenge of detecting extremely short FOG episodes in the Mobilise-D cohort, which primarily consists of mild freezers with the 360° turning-in-place task. Fine-tuning allowed the model to capture some of these brief episodes; however, the improvements were not consistent.

**Fig. 3:**
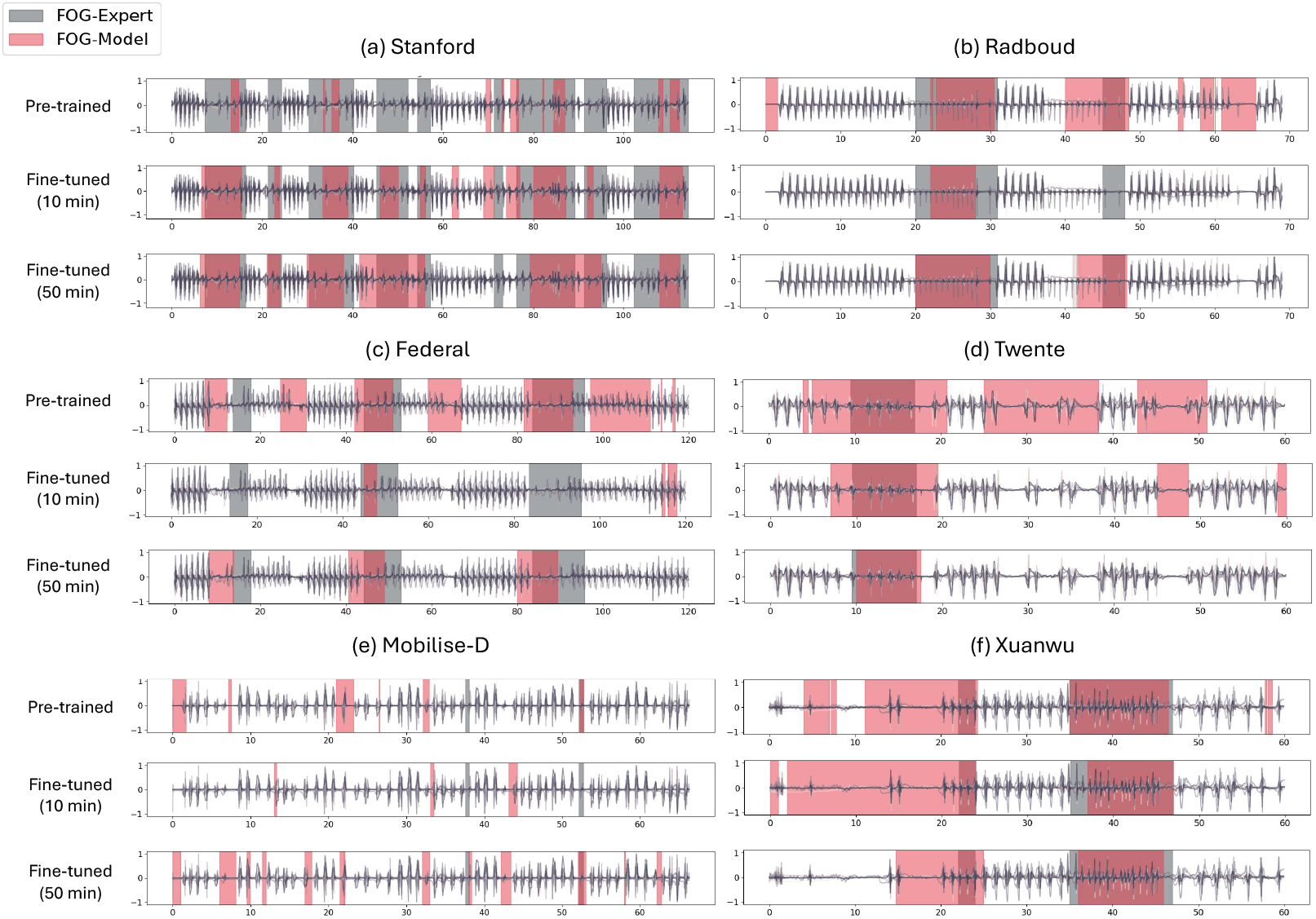
Model predictions (pre-trained, fine-tuned with 10 minutes of data, and fine-tuned with 50 minutes of data) versus expert annotations across all external cohorts. Normalized IMU signals for each trial were shown: with right foot for Stanford, Mobilise-D, and Radboud; left foot for Federal; right ankle for Twente; and right tibia for the two-IMU Xuanwu dataset.

#### 2.3.1 Understanding dataset-specific differences

As discussed in previous sections, the performance drop observed when applying the pre-trained model to external datasets highlights the presence of cohort differences. While some differences, such as variations in IMU configurations (e.g., using five IMUs versus one or two IMU), are apparent, others are more subtle. In this subsection, we examine three less obvious but critical cohort differences: FOG-provoking task (i.e., Radboud and Stanford datasets), FOG annotation criteria (i.e., Stanford dataset), and variations in patient spectrum bias (i.e., Mobilise-D dataset).

In the Radboud dataset, the pre-trained model frequently misclassified volitional stopping periods as FOG episodes, contributing to higher FP rates (Figure 4a). To explore an intervention beyond model fine-tuning, we simulated an “expert-review” workflow in which an expert manually removed all FPs from non-FOG trials (a potential feature of the AID-FOG platform). This correction markedly improved agreement with ground truth annotations, as shown in Table 2, increasing ICC(%TF) from 0.481 to 0.893 and ICC(#FOG) from 0.098 to 0.642. These findings highlight the value of expert refinement, particularly when task-specific information (e.g., stopping periods) is documented during data collection.

**Fig. 4:**
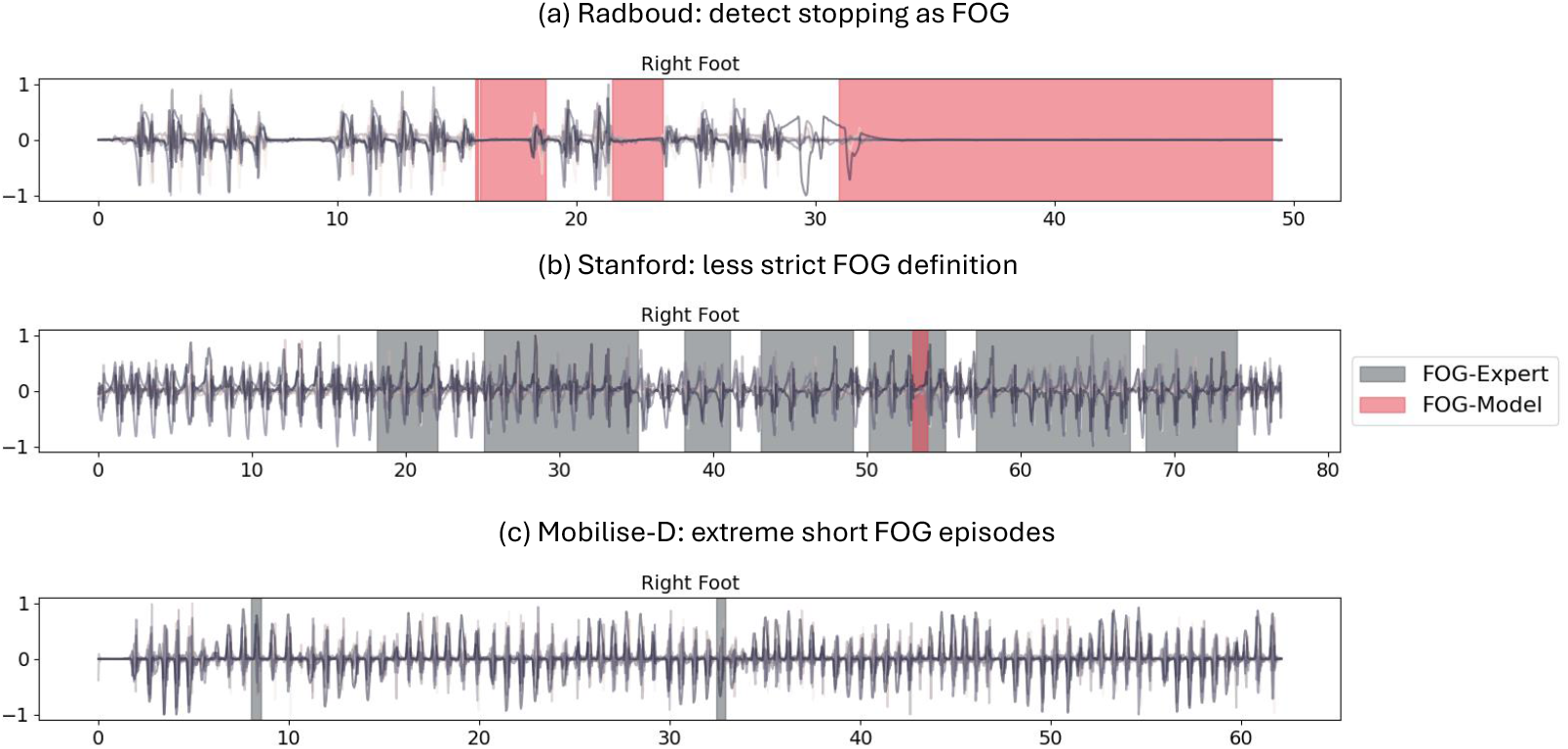
Pre-trained model predictions (red) versus expert annotations (gray) for (a) the Radboud cohort, showing difficulty in distinguishing stopping from FOG, (b) the Stanford cohort, showing difficulty in detecting shuffling FOG episodes, and (c) the Mobilise-D cohort, showing the difficulty in identifying extremely short FOG episodes in mild freezers. Normalized IMU signals from the right foot are displayed.

In the Stanford cohort, challenges arose from differences in FOG annotation criteria. As illustrated in Figure 4b, the Stanford dataset applied a labeling scheme that included prolonged shuffling episodes with observable IMU motion as FOG. These instances were not initially detected by the pre-trained model, which had been developed using a different annotation protocol. Fine-tuning allowed the model to adapt to this expanded definition, but also led to an increased #FPs (Figure 3a). This discrepancy reflects a limitation in the original FOG definition Nutt et al (2011), which allowed for subjective interpretation during annotation. This challenge was also emphasized by Mancini et al (2025b) and motivated efforts to establish a standardized definition of FOG Lewis et al (2022).

The Mobilise-D dataset presented a different challenge related to patient spectrum bias. As shown in Figure 4c, many annotated FOG episodes were extremely brief, making them difficult to detect from the models. To assess annotation reliability, a third independent rater annotated 29 Mobilise-D trials, primarily from the Kiel and Tel Aviv center, to better reflect clinical diversity. The ICC between the fine-tuned model and the original ground truth was 0.540 for %TF and 0.756 for #FOG, whereas the ICC between the third rater and the ground truth was 0.757 and 0.543, respectively. These findings suggest that FOG detection in the Mobilise-D dataset is intrinsically challenging, even for experienced human raters. Visual comparisons of the model predictions (fine-tuned with 50 minutes of data), the original annotations, and the third rater’s annotations of the 29 trials are provided in Figures E2 and E3 (in Supplementary Appendix E).

## 3 Discussion

In this study, we developed and externally validated a DLA for assessing FOG severity using IMU data from a large cohort of people with PD (PwPD) across six independent datasets. Our results showed that while the DLA model achieved strong agreement on the local KU Leuven cohort for %TF (ICC = 0.886, CI=[0.79, 0.90]) and fair agreement for #FOG (ICC = 0.573, CI=[0.28, 0.71]), its performance dropped on the external datasets, averaging fair agreement for %TF (ICC = 0.562 *±* 0.141, CI=[−0.01, 0.78]) and poor agreement for #FOG (ICC = 0.298 *±* 0.270, CI=[−0.16, 0.49]).

To address this generalization gap, we explored fine-tuning with expert-annotated external data across varying durations. Our analysis demonstrated that fine-tuning the model reached a plateau for both ICC(%TF) and ICC(#FOG) using 50 minutes of annotated data for training, with improved ICC(%TF) to 0.732 *±* 0.138 (+30.2%) and ICC(#FOG) to 0.623 *±* 0.214 (+108.9%). In contrast, retraining models from scratch reached a plateau with more data (90 minutes for ICC(%TF) of 0.708 *±* 0.195 and 270 minutes for ICC(#FOG) of 0.528 *±* 0.244). These findings highlight the data efficiency and performance benefits of fine-tuning, which enables better generalization with less annotated data.

However, fine-tuning alone cannot replace expert review at the current stage, since our best ICCs still lie at the lower boundary of reported inter-rater ranges (0.730-0.990 for %TF and 0.390-0.950 for #FOG D’Cruz et al (2021); Morris et al (2012); O’Day et al (2022); Mancini et al (2021); Kondo et al (2022)). As a result, we consider that fully automated FOG detection is not yet feasible due to the lack of unified standards and the resulting variability across studies such as sensor configurations, task design, and annotation criteria. While unified standards are being developed Mancini et al (2025b); Lewis et al (2022); Mancini et al (2025a), progressive model improvement with fine-tuning can already provide a temporary solution to make the annotation workload more efficient, since only a subset of data (e.g., 50 minutes) requires manual annotation. In our proposed AI-assisted workflow (Figure 1c and Appendix H), experts would first review model-generated annotations to (1) flag any missed FOG episodes and (2) remove clear FPs. Those corrections could be further used to fine-tune the model, shifting its decision boundary to catching previously missed FOG episodes and dropping FPs. A practical use case would imply uploading pilot data to generate customized models for clinical trials. Despite these advantages in our study, we fine-tuned solely on existing ground-truth labels and did not incorporate expert-adjusted annotations; in practice, initial exposure to model predictions may bias experts toward correcting only the most obvious errors. Future user studies should evaluate whether fine-tuning the model using these expert-refined annotations could reproduce or even improve the current performance gains and quantify any annotation bias introduced by the selective correction.

In this study, we assessed both fine-tuning and training-from-scratch using 7-fold participant-level cross-validation: holding out one fold for testing each run and averaging results to avoid split-dependent effects. Within each training fold, we built nested annotation sets of increasing duration (10, 20, …, 360 min) by sequentially adding participants while preserving the cohort’s overall %TF distribution. This design ensures that performance gains reflect added annotation time rather than changes in participant mix or FOG prevalence, and mirrors the expert-in-the-loop workflow of incrementally uploading extra minutes from the same patients. However, note that cohorts differed in their recording lengths per participant. For example, with a 50-minute fine-tuning, Stanford which contains more trials per participant could include five 10-minute segments per participant with a total of five participants; whereas Mobilise-D can be assembled from 50 one-minute trials across 50 participants. Although this does not imply how many participants must be recruited or re-annotated, our approach quantifies the exact minutes of data needed to reach any target performance level. Although annotating 50 minutes of data still requires effort, it is considerably less burdensome than annotating the entire cohort.

Additionally, detecting FOG in the predominantly mild-freezer Mobilise-D cohort demonstrated to be particularly challenging, illustrating the risks of applying models trained on severe freezers to mildly affected populations. Although fine-tuning can increase performance in this group, the improvements are smaller and the ceiling remains lower than in severe-freezer cohorts, making expert review of annotations necessary. Until sufficiently large datasets of mild and pre-freezers exist, expert reviews are essential, and an AI-assisted workflow could potentially benefit this process.

Compared to other large-scale efforts, Salomon *et al*. ran a machine-learning challenge using over 90 h of daily-living accelerometer data from a single IMU, training on 833 trials from 62 participants and 91 trials from 38 participants with PD, with public (945 trials, 26 participants) and non-public test sets (391 trials, 14 participants) Salomon et al (2024). The top five DLAs all incorporated self-attention or transformer blocks, consistent with our finding that LTContext, with attention modules, outperforms MS-TCN and ASFormer in FOG detection on external datasets. The top-performing model in their study achieved a recall of 0.78 and a specificity of 0.94, with a strong agreement for %TF (ICC *>* 0.87) and fair agreement for #FOG (ICC *>* 0.50). These results exceed those we observed when evaluating our models on the external cohorts. However, it is important to note that although their data came from three centers (KU Leuven, Harvard, and Tel Aviv), their train–test split minimized cohort heterogeneity, such as differences in task protocols, sensor configurations, and annotation criteria between training and evaluation sets. Borzì *et al*. evaluated a single-ankle IMU model on 22 participants (101 FOG episodes) and externally validated it on two independent datasets (545 episodes), achieving 88-95% detection sensitivity but suffering from high FP rates Borzì et al (2025). Similarly, in our internal–to–external validation, we observed an increase in #FPs on external datasets. However, after fine-tuning, the #FPs decreased, and both specificity and precision improve (see Supplementary Appendix G).

As the field awaits updated FOG definitions Lewis et al (2022), re-annotating existing datasets to align with new standards would demand efforts. However, as this update represents a shift in annotation criteria, similar to the differences we observed in the Stanford cohort, our fine-tuning approach can adapt pre-trained models to future definitions without retraining models from scratch. Similarly, fine-tuning could possibly streamline this process by progressively improving model adaptation to future definitions. Ultimately, the ICFOG proposed guidelines on standardizing FOG annotation criteria Lewis et al (2022), IMU sensor protocols, and FOG-provoking tasks Mancini et al (2025b) could help reduce inter-center variability, enable model external performance to approach internal validation performance, and potentially remove the need for fine-tuning. Until such consensus and standardization are in place, model fine-tuning remains a temporary solution to make the annotation workload more efficient.

In conclusion, we presented a DLA trained on a large local cohort within KU Leuven and externally validated the model on six external cohorts of people with PD and FOG. Although performance dropped due to cohort differences, fine-tuning with limited external data (e.g., 50 minutes) substantially improved generalizability, albeit not yet to optimal levels, underscoring the continued need for expert review. To support both trust and model adaptation, we proposed an AI-assisted, expert-in-the-loop workflow: the DLA suggests FOG episodes, experts correct and refine these annotations, and the model is iteratively fine-tuned. Finally, we are building the AID-FOG platform, an interactive platform tailored for clinical annotation and model updating. Together, these contributions represent important steps toward making AI-assisted FOG analysis in PD more robust, clinically meaningful, and scalable across diverse real-world settings.

## 4 Methods

### 4.1 Study design and participants

This multicentre retrospective study developed a DLA, trained on an local cohort (KU Leuven) and externally validated on six cohorts. Data were collected from eight universities: KU Leuven (non-public), Radboud University (non-public) Janssen et al (2017), Stanford University (public) O’Day et al (2022), Federal University of ABC (public) Ribeiro De Souza et al (2022), University of Twente (public) Delgado-Terán (2025); Delgado-Terán et al (2025), Beijing Xuanwu Hospital Zhang et al (2022); Li (2021), and the Mobilise-D project (non-public) Goris et al (2025), which included data from KU Leuven, University Medical Center Schleswig-Holstein Campus Kiel, and Tel Aviv Sourasky Medical Center. We included only public datasets that provide lower-limb IMUs with both accelerometer and gyroscope data collected during controlled tasks; datasets offering only accelerometers Salomon et al (2024); Bachlin et al (2009) or only upper-limb sensors Rodríguez-Martín et al (2017) were excluded.

- **KU Leuven Cohort (Local Dataset)**: Included total of 85 PwPD with FOG across four datasets. IMU configurations and FOG-provoking tasks varied across datasets:
  - Dataset 1: Included 40 PwPD (30 self-reported freezers, 10 non-freezers) Yang et al (2024), with 2 participants excluded due to technical failure. Tasks: Timed Up and Go (TUG) test and one-minute alternating 360° turns under both OFF (*>*12 h since last dose) and ON (*≥* 1 h post-intake) medication states, each performed with and without a cognitive dual task (i.e., an auditory Stroop task Kestens et al (2021)) and with and without a volitional stopping period between tasks. Used five Shimmer3 IMUs (64 Hz, Shimmer Research, Ireland) placed on the pelvis and both sides of the tibia and talus.
  - Dataset 2: Included 28 PwPD (14 freezers, 14 non-freezers) Spildooren et al (2010), with 1 excluded due to technical issue. Tasks: 5m straight-line walk with/without 180°/360° turns under OFF-medication. Used synthesized IMUs virtually placed at the same positions as dataset 1, derived from Vicon motion capture (100 Hz, 8-camera system, Vicon, UK) Ghorbani and Black (2021); Yi et al (2021b).
  - Dataset 3: Included 8 PwPD with baseline and 12-month follow-up data (out of 17) Vervoort et al (2016); all trials from both visits for each participant were combined (not entered as separate participants). Tasks: straight walking and 360° turns under OFF-medication. Used synthesized IMUs virtually placed at the same positions as dataset 1, derived from Vicon motion capture (100 Hz, 8-camera system, Vicon, UK) Ghorbani and Black (2021); Yi et al (2021b).
  - Dataset 4: Included 12 PwPD with a walking/slalom task (three walking/slalom bouts separated by stops) under ON-medication, each completed with and without a cognitive dual task. Used five APDM IMUs (128 Hz, APDM, USA) placed as dataset 1. Note that this dataset originates from an ongoing study (ClinicalTrials.gov NCT05511597).
- **Radboud University Cohort (External Dataset 1)** Janssen et al (2017): Included 25 PwPD with FOG. Used 17 Xsens Awinda IMUs (60 Hz, Xsens Technologies, Netherlands). IMUs covered full-body motion capture. Tasks: 15m walking with volitional stopping, walking straight, and auditory-cued 360° turns. Conducted under OFF-medication.
- **Stanford University Cohort (External Dataset 2)** O’Day et al (2022): Included 7 PwPD with FOG, each completing 5-14 repetitions of an OFF-medication slalom course (two ellipses and two figure-eight turns around barriers) while wearing seven APDM IMUs (128 Hz, APDM, USA) at the chest, lumbar spine (pelvis), ankles (tibia), and feet (talus); data from multiple visits (up to 44 months apart) were aggregated per participant rather than treated as separate participants.
- **Federal University of ABC Cohort (External Dataset 3)** Ribeiro De Souza et al (2022): Included 35 PwPD with FOG. Used 1 Physilog 5 IMU (128 Hz, Gait Up, Switzerland) placed on the left lower leg (tibia position). Task: two-minute alternating 360° turn. Conducted under ON-medication.
- **University of Twente Cohort (External Dataset 4)** Delgado-Terán (2025); Delgado-Terán et al (2025): Included 21 PwPD with FOG. Used 1 Movisens Move 4 IMU (60 Hz, movisens GmbH, Germany) mounted on the right ankle (tibia position). Data were recorded in semi–free-living conditions, encompassing daily routines (e.g. cleaning, setting the table, mopping, making the bed), outdoor walking, and a clinical assessment. The full dataset was divided into “all movements” and “walking-and-turning” segments; this study analyzed only the walking-and-turning portion. All recordings were performed under OFF-medication.
- **Beijing Xuanwu Hospital (External Dataset 5)** Zhang et al (2022); Li (2021): Included 12 PwPD with FOG. A single TDK MPU6050 IMU (100 Hz, TDK InvenSense, USA) was mounted on the left or right tibia (or both). Two walking-and-turning tasks were performed: Participants stood from a chair, walked 5 m into an 18 m corridor, navigated three obstacles by rotating around them, made a 180° turn at the far end, then retraced their steps back to the chair and sat down; Participants walked 3 m from the chair to a 0.6 m square, executed a 180° turn inside the square, and returned to sit. All trials were conducted under OFF-medication. Both raw and filtered data are available in this public dataset; since the raw data include signals recorded during patient preparation, we used only the filtered data in this study. Detailed processing steps are described in the original publications.
- **Mobilise-D Project Cohort (External Dataset 6)** Goris et al (2025): Included 52 PwPD from KU Leuven, 90 PwPD from University Medical Center Schleswig-Holstein Campus Kiel, and 35 PwPD from Tel Aviv Sourasky Medical Center (after excluding 6, 9, and 6 participants respectively for technical issues). Used APDM IMUs (128 Hz; APDM, USA) at the pelvis and both sides of the tibia and talus. Task: one-minute alternating 360° turns with a concurrent cognitive dual task. Conducted under ON-medication. Note: to represent mild-to-moderate freezers, the KU Leuven subset was merged with the Kiel and Tel Aviv cohorts into a single external cohort (termed Mobilise-D).

All cohorts included synchronized video recordings for FOG annotation. The KU Leuven datasets were annotated by two external raters using the criteria: “FOG was defined as a brief episode with the inability to produce effective steps” Nutt et al (2011). Specifically, an episode started only when the participant’s foot suddenly ceased producing an effective forward step and displayed FOG-related features, and ended only after at least two effective steps had been completed (these two steps were not included in the episode) Gilat et al (2018).

The Stanford dataset was annotated by two experienced raters using the criteria: “a FOG event was defined by loss of alternating stepping, complete cessation of forward motion, or trembling of the legs” Heremans et al (2013).

The Federal dataset was annotated by two experienced raters using the criteria: “The beginning of a FOG episode was defined when the turn pattern (alternating right and left steps) was arrested, or when participants appeared to be trying unsuccessfully to initiate or continue the turn. The end of the episode was marked by the execution of an effective step followed by continuous turning” Mancini et al (2021).

The Mobilise-D dataset was annotated by two independent expert raters and verified by a third experienced moderator in cases of disagreement. Each FOG episode was further labeled as either predominantly “trembling”, characterized by oscillatory movements between 3-8 Hz visible in the legs for more than 50% of the episode, or predominantly “akinesia”, defined by an absence of visible leg trembling and a lack of foot progression for more than 50% of the episode.

The Radboud and Twente cohorts were annotated by two experienced raters following the clinical definition provided by Nutt et al (2011): “FOG was defined as a brief, episodic absence or marked reduction of forward progression of the feet despite the intention to walk.” The Xuanwu cohort was annotated by two physicians Zhang et al (2022), although the specific criteria used to label FOG episodes were not reported.

Clinical assessments were available in all cohorts except Stanford, including:

- Movement Disorders Society Unified Parkinson’s Disease Rating Scale (MDS-UPDRS III) Goetz et al (2008)
- New Freezing of Gait Questionnaire (NFOG-Q) Nieuwboer et al (2009) (Xuanwu cohort included the Freezing of Gait Questionnaire (FOGQ) instead Giladi et al (2000)) Some cohorts also included:
- Hoehn & Yahr (H&Y) Scale Hoehn and Yahr (1967)
- Mini-Mental State Examination (MMSE) Folstein et al (1975)
- Montreal Cognitive Assessment (MoCA) Nasreddine et al (2005)

### 4.2 Deep learning algorithm

#### 4.2.1 Data harmonization

To harmonize the datasets, we first applied a zero-phase fourth-order Butterworth low-pass filter with a 16 Hz cutoff frequency, selected to preserve the locomotor (0-3 Hz) and freezing (3-8 Hz) bands Moore et al (2008). All IMU data were then resampled to a uniform 32 Hz sampling frequency. Further preprocessing was performed using the `gaitmap` Python library Küderle et al (2024), which provides automated IMU signal processing tools. While its algorithms are primarily designed for foot-worn IMUs, they were used to ensure consistent sensor alignment across datasets with similar placements.

First, `gaitmap`‘s gravity alignment algorithm was applied, rotating signals so that the gravitational component is fully captured by the z-axis. A value of 9.81 was then subtracted to center all signals around zero. This step was skipped for datasets where the gravitational component was already removed. Next, IMU signals were aligned using `gaitmap`‘s PCA-based algorithm, which detects the medio-lateral component as the first principal component and rotates the signals accordingly. Since PCA sign orientation depends on data distribution, an additional correction was applied to prevent a 180° misalignment of accelerations. This final correction was performed using `gaitmap`‘s trajectory reconstruction algorithm. Lastly, the standard offset was removed by subtracting the mean of each signal before feeding the data into the model.

As most datasets included five IMUs, adaptations were required for the Federal, Twente, and Xuanwu datasets, which each employed either one or two IMUs. To maintain consistent input dimensions across all datasets, missing IMU channels for these three dataset were filled with zeros during preprocessing.

The Twente dataset underwent preprocessing as part of its original public release, where signals were segmented into 120 time-step windows with an 87.5% overlap for FOG segments and a 50% overlap for non-FOG segments. To ensure consistency with the other datasets, the segmented windows were reorganized into a continuous sequence for analysis. Unlike other datasets, where tasks lasted less than 2 minutes, the Twente dataset included longer semi-free-living walking and turning sequences. For consistency in processing across cohorts and prevent memory overload, these extended trials were partitioned into 2-minute segments before model training.

#### 4.2.2 Problem statement

Formally, the IMU data of each gait task can be represented as a 2D matrix 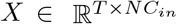, where *T* represents the number of samples (duration of the gait task), *N* the number of IMUs, and *C*_*in*_ the number of features per IMU (3-axis accelerometer and 3-axis gyroscope). Similarly, the ground truth can be represented as a 2D matrix *Y ∈* ℝ^*T* × *L*^, where *T* represents the number of samples and *L* the number of classes (i.e., *L* = 2 for FOG and non-FOG).

#### 4.2.3 Deep neural network design

Fine-grained FOG detection often suffers from over-segmentation, i.e., splitting true FOG episodes into multiple short segments, which complicates expert review in AI-assisted annotation tools such as the AID-FOG platform. MS-TCN mitigates this issue by stacking TCN stages with a smoothing loss to enforce temporal consistency Farha and Gall (2019); Filtjens et al (2023). Building on MS-TCN, ASFormer Yi et al (2021a) and LTContext Bahrami et al (2023) integrated attention modules (recently shown to increase FOG detection performance Salomon et al (2024)) to capture long-range dependencies and further refine action segmentation. To determine which refinement-based sequence model performs best across external cohorts for FOG detection, we benchmarked MS-TCN, ASFormer, and LTContext on all external datasets. A hyperparameter analysis (Supplementary Appendix B) and model selection study (Supplementary Appendix C) identified LTContext as the best-performing model for external datasets. The LTContext architecture is detailed below, while descriptions of MS-TCN and ASFormer are provided in Supplementary Appendix A.

The input sequence of IMU features, 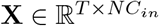, first passes through an input projection layer (a 1 *×* 1 convolution), mapping the original input features to an internal hidden representation of shape ℝ^*T* × *C*^, where *T* is the sequence length and *C* is the number of hidden features. This transformed representation is then processed by the initial prediction stage, consisting of 10 LTContext blocks. Each LTContext block integrates:

- A dilated TCN layer with exponentially increasing dilation factors to expand the receptive field.
- A Gaussian Error Linear Unit (GeLU) activation function for non-linearity.
- Instance Normalization to stabilize training.
- Windowed context attention, which partitions the sequence into small non-overlapping windows and computes attention within each window to capture short-term dependencies.
- Long-term (global) attention, which sparsely samples elements across the entire sequence to model long-range dependencies.

After processing through these 10 LTContext blocks, the output remains ℝ^*T* × *C*^. A linear projection layer then reduces the feature dimensionality to ℝ^*T* × *C/*2^, followed by a SoftMax layer that produces an initial set of class probabilities **Ŷ** ^(0)^ *∈* ℝ^*T* × *L*^.

The predictions are refined through three additional stages, each containing LTContext blocks. Unlike the initial stage, these blocks replace the input feature sequence with the predictions from the previous stage. At each refinement stage *s*, the model generates updated predictions **Ŷ** ^(*s*)^ *∈* ℝ^*T ×L*^.

#### 4.2.4 Loss function

A loss function (ℒ) computed the sum of the losses over all *N* stages:

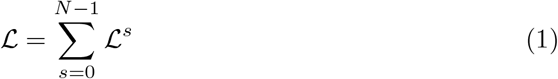

where ℒ_*s*_ represents the loss for the output probabilities at stage *s*. Each stage’s loss combines the cross-entropy loss (*L*_*cls*_) and the smoothing loss (*L*_*T* −*MSE*_), with the latter weighted by a hyperparameter *λ*_*T* −*MSE*_ that controls its contribution. In this study, *λ*_*T* −*MSE*_ is set to 0.15, as recommended in previous research Farha and Gall (2019).

The loss for each stage (*L*^*s*^) is formalized as:

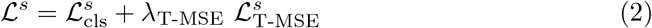

The cross-entropy loss measures classification performance and is defined as:

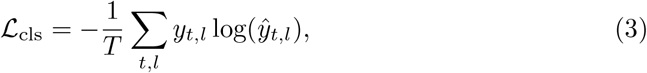

where *T* is the sequence length, *L* is the total number of classes, *y*_*t,l*_ is the ground truth for class *l* at time step *t*, and *ŷ*_*t,l*_ is the predicted probability of class *l* at time *t*. The smoothing loss, ℒ_T-MSE_, is a truncated mean squared error applied to the log-probabilities of consecutive samples:

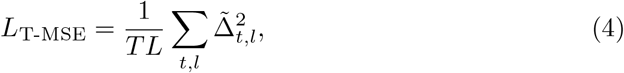

where 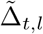 is:

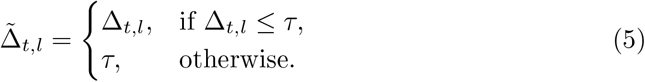

Here, Δ_*t,l*_ represents the absolute difference in log-probabilities between consecutive time steps:

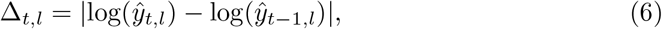

and *τ* is a hyperparameter defining the threshold for truncating the smoothing loss.

### 4.3 Evaluation and training strategies

Supplementary Appendix B details the selection of key hyperparameters for the LTContext model. The final configuration included a batch size of 1, 10 layers, 4 stages, and 64 hidden features, while other hyperparameters remained as specified in the original LTContext publication Bahrami et al (2023).

The model was first pre-trained on the KU Leuven cohort using the Adam optimizer (*β*_1_ = 0.9, *β*_2_ = 0.999) Kingma and Ba (2015) for 50 epochs. Training began with a learning rate of 0.00025, which was gradually reduced to 0.00005 using a cosine decay schedule Bahrami et al (2023).

We evaluated two strategies for adapting the model to external cohorts: (1) training from scratch and (2) fine-tuning the pre-trained model. Both strategies used 7-fold participant-level cross-validation. In each fold, we sampled training durations in 10-minute increments up to 60 min (to match Stanford’s maximum) and in larger steps thereafter (90, 120, 150, …, 360 min), as a compromise between temporal resolution and computational feasibility. This setup required 7 folds *×* 5 cohorts *×* 2 strategies *×* 16 time points = 1120 separate model trainings. The maximum available durations varied by cohort (Stanford: 60 min; Mobilise-D: 150 min; Federal: 120 min; Radboud and Twente: 360 min). For reporting averaged results, we used each cohort’s last available time point, assuming performance generally improves or plateaus with more data Zhu et al (2016). We defined the plateau as the smallest training duration beyond which additional data yields no more than a one-percentage-point gain in the target metric (e.g., ICC(%TF)) Viering and Loog (2022).

For models trained from scratch, all hyperparameters and optimization settings mirrored those of pre-training, including training for 50 epochs using Adam and cosine learning rate decay.

Fine-tuning followed a selective learning rate strategy inspired by Zhu et al (2023). The input projection and LTContext blocks (feature extraction layers) were fine-tuned with a learning rate of 0.00025, while the prediction layers (output projections and refinement stages) were updated with a higher learning rate of 0.0025 to accelerate adaptation. Optimization used the Adam optimizer (batch size = 1) with cosine decay to 0.00005. As described in Supplementary Appendix D, the number of backpropagation steps was fixed at 3000 for all training durations, based on convergence analysis using the 360-minute condition. All other hyperparameters matched the pre-training configuration.

### 4.4 Metrics and statistics

The primary metric for evaluating action segmentation reliability was the sample-wise F1 score. To assess temporal consistency, we additionally used the segment-wise F1 score with a 50% intersection-over-union threshold (F1@50) Lea et al (2017), which penalizes over-segmentation by requiring predicted segments to closely align with ground truth annotations Filtjens et al (2022). The segment-wise F1@50 is particularly relevant for the proposed AID-FOG platform, as lower F1@50 scores imply that predicted FOG episodes are temporally misaligned, requiring more manual adjustments by clinicians to refine segment boundaries. To ensure a detailed and unbiased evaluation of model performance, we reported metrics separately for FOG and non-FOG trials. For trials containing expert-annotated FOG episodes (FOG trials), both the sample-wise F1 and segment-wise F1@50 were computed. For non-FOG trials, performance was assessed using the following metrics:

- Mean number of false positive (FP) episodes per trial (#FP per trial)
- Mean FP duration per trial

Lower values for both #FP per trial and mean FP duration indicate higher precision, reflecting fewer and shorter erroneous FOG detections. These metrics are also practically meaningful for the AID-FOG platform, as more FPs translate to more manual clicks and corrections by annotators. Figure 5 illustrates the computation of these four evaluation metrics. To align with prior FOG detection research Mancini et al (2025b), we also computed the traditional machine learning metrics, e.g., accuracy, recall, specificity, and precision, across the whole dataset of each external cohort. The results of these metrics are detailed in Supplementary Appendix G.

**Fig. 5:**
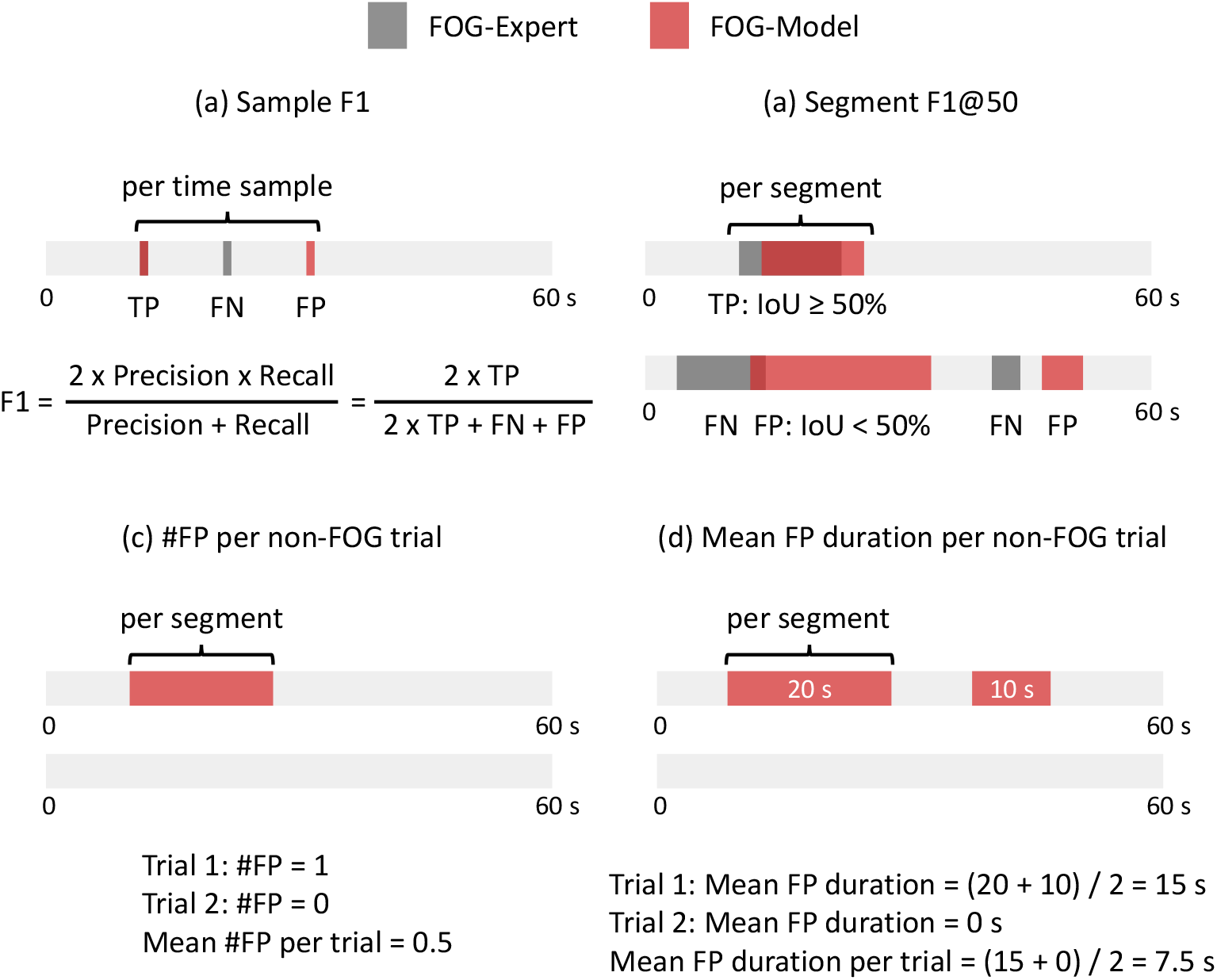
An illustration of the calculation of sample-wise F1 and segment-wise F1@50 scores for FOG trials, and the computation of the mean #FP per trial and mean FP duration per trial for non-FOG trials.

In this study, FOG severity was quantified using the number of FOG episodes (#FOG) and the percentage time spent frozen (%TF), defined as the cumulative duration of all FOG episodes divided by the total duration of the gait task Morris et al (2012). To assess reliability in FOG severity estimation, we computed the intra-class correlation coefficient (ICC) using a two-way random effects model (ICC(2,1)), reflecting absolute agreement for a single measurement.

All metrics were computed at the participant level by aggregating all gait tasks per participant. The statistical significance of F1-score differences between original annotations and an additional rater, as well as those between original annotations and model predictions, was assessed using the paired Student’s t-test. Similarly, paired Student’s t-tests were applied to compare different models, with multiple comparisons adjusted using the Li correction (David). F-tests were used to evaluate ICC significance McGraw and Wong (1996). Homogeneity of variances was tested using Levene’s test, and normality was verified using the Shapiro-Wilk test. Whenever these assumptions were violated, we instead applied the nonparametric Wilcoxon signed-rank test. The significance level for all statistical tests was set at 0.05.

## Data availability

The local cohort datasets in KU Leuven and external Radboud dataset analyzed in this study are not publicly available due to restrictions on sharing sensitive patient health information. In contrast, the following external datasets are publicly accessible:

- Stanford O’Day et al (2022): Available at https://github.com/stanfordnmbl/imu-fog-detection
- Twente Delgado-Terán (2025); Delgado-Terán et al (2025): Available at https://data.4tu.nl/datasets/40e06061-f441-43b5-9235-006829206509
- Federal Ribeiro De Souza et al (2022): Available at https://figshare.com/articles/dataset/14984667
- Xuanwu Zhang et al (2022): Available at https://data.mendeley.com/datasets/r8gmbtv7w2/3

## Code availability

The DL architectures were implemented in PyTorch 2.2.2+cu118 Paszke et al (2019), leveraging publicly available repositories for MS-TCN Farha and Gall (2019),

ASFormer Yi et al (2021a), and LTContext Bahrami et al (2023). Statistical analyses and visualizations were conducted using SciPy 1.7.11, bioinfokit 2.1.0, statsmodels 0.13.2, and pingouin 0.3.12 for statistical computations, and matplotlib 3.8.4 for plotting. The scripts were written in Python 3.10.14, with scamamp Calvo and Santafé (2016) implemented in R for additional statistical modeling. The code developed for this study is not publicly available, but may be provided upon reasonable request.

## Acknowledgments

This study was funded, in part, by the KU Leuven Industrial Research Fund (C3/20/109), the AidWear project funded by the Federal Public Service for Policy and Support, and the AID-FOG project by the Michael J. Fox Foundation for Parkinson’s Research under Grant ID: MJFF-024628. This paper presents independent research supported by the NIHR Newcastle Biomedical Research Centre (BRC), UK. The NIHR Newcastle BRC is a partnership between Newcastle Hospitals NHS Foundation Trust, Newcastle University, and Cumbria, Northumberland and Tyne and Wear NHS Foundation Trust and is funded by the National Institute for Health and Care Research (NIHR). PY was supported by the Ministry of Education (KU Leuven - Taiwan) scholarship. JC was supported by the strategic basic research project RevalExo (S001024N) funded by the Research Foundation Flanders and the Flanders AI Research Program. MG was supported by the Research Foundation Flanders (1SHEK24N). Computational resources were provided by the VSC (Flemish Supercomputer Center), funded by the Research Foundation - Flanders (FWO) and the Flemish Government.

Additionally, the Mobilise-D project has received funding from the Innovative Medicines Initiative 2 Joint Undertaking under grant agreement no. 820820. This Joint Undertaking receives support from the European Union’s Horizon 2020 research and innovation program and the European Federation of Pharmaceutical Industries and Associations (EFPIA). This publication reflects the authors’ views and neither IMI nor the European Union, EFPIA, or any Associated Partners are responsible for any use that may be made of the information contained herein.

## Author contributions

Study design: BF, PG, MG, AN, and BV. Data analysis: BF, JC, and PY. Neural network design and implementation: JC and PY. Data collection and preparation: EK, JN, RW, LA, AY, LR, CH, CS, WM, DB, MB, JH, MG, MG, PG, and AN. Manuscript drafting: BF and PY. All authors provided feedback on manuscript revisions and approved the final version.

## Ethics approval and consent to participate

All ten retrospective datasets (four from KU Leuven and six external cohorts) were originally collected under approvals from their local ethics committees at the time of data collection; reuse of these data was approved by the UZ/KU Leuven Ethics Committee (S65059). All participants had provided written informed consent. The user-study with three clinical experts to develop AID-FOG received approval from the Social and Medical Ethics Committee (SMEC; ref. G-2023-7462), and all experts also gave written informed consent.

## Appendix A Deep learning architectures

We compared three state-of-the-art sequence learning architectures for FOG detection: MS-TCN, ASFormer, and LTContext. ASFormer and LTContext were developed as extensions of the MS-TCN framework. This section outlines the architecture of these models.

All models first apply an input projection layer (1 *×* 1 convolution) to map input features to an internal network representation:

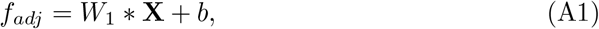

where *f*_*adj*_ *∈* ℝ^*T* × *C*^ is the adjusted feature map, 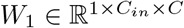 are the convolution weights, and *b ∈* ℝ^*C*^ is the bias term.

For MS-TCN, the internal network representation is first processed through a single-stage TCN consisting of 10 ResNet-style TCN layers. Each layer employs a kernel size of 3, with exponentially increasing dilation rates: {1, 2, 4, 8, 16, 32, 64, 128, 256, 512}. The output of the single-stage TCN is passed through a SoftMax layer, implemented as a 1 *×* 1 convolutional layer, to generate initial class probabilities. These probabilities are then refined through three additional multi-stage TCN layers, each mirroring the architecture of the single-stage TCN.

For ASFormer, the internal network representation is processed through an Encoder consisting of 10 encoder blocks, each comprising a dilated TCN layer, a ReLU activation function, Instance Normalization, a Self-Attention layer, and a 1 *×* 1 convolutional layer. The Encoder generates initial class probabilities through a SoftMax layer. These probabilities are refined by three additional Decoder stages, each containing 10 decoder blocks. Each decoder block includes a dilated TCN layer, a ReLU activation function, Instance Normalization, a Cross-Attention layer (attending to encoder features), and a 1 *×* 1 convolutional layer.

For LTContext, the internal network representation is processed through an initial stage consisting of 10 LTContext blocks. Each block integrates a dilated TCN layer with exponentially increasing dilation factors, a Gaussian Error Linear Unit (GeLU) activation function, Instance Normalization, windowed context attention, and long-term context attention. Both attention mechanisms use the input features as queries and keys. After this stage, a linear layer reduces the feature dimensionality to half of the original size *C*, and the output is passed through a SoftMax layer to generate initial class probabilities. The predictions are further refined across three additional stages, where each LTContext block applies attention mechanisms to the model’s predictions from the previous stage instead of the original input features.

For all three models, predictions from each stage are concatenated and used to compute the final loss. The SoftMax output layer, applied at each stage, is formulated as:

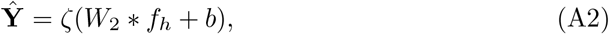

where **Ŷ** *∈* ℝ^*T* × *L*^ are the predicted class probabilities, *f*_*h*_ *∈* ℝ^*T* × *C*^ is the final feature representation, *W*_2_ *∈* ℝ^1*×C×L*^ are the convolution weights, and *ζ* is the SoftMax function.

A visual comparison of the three model architectures is presented in Figure A1.

**Fig. A1:**
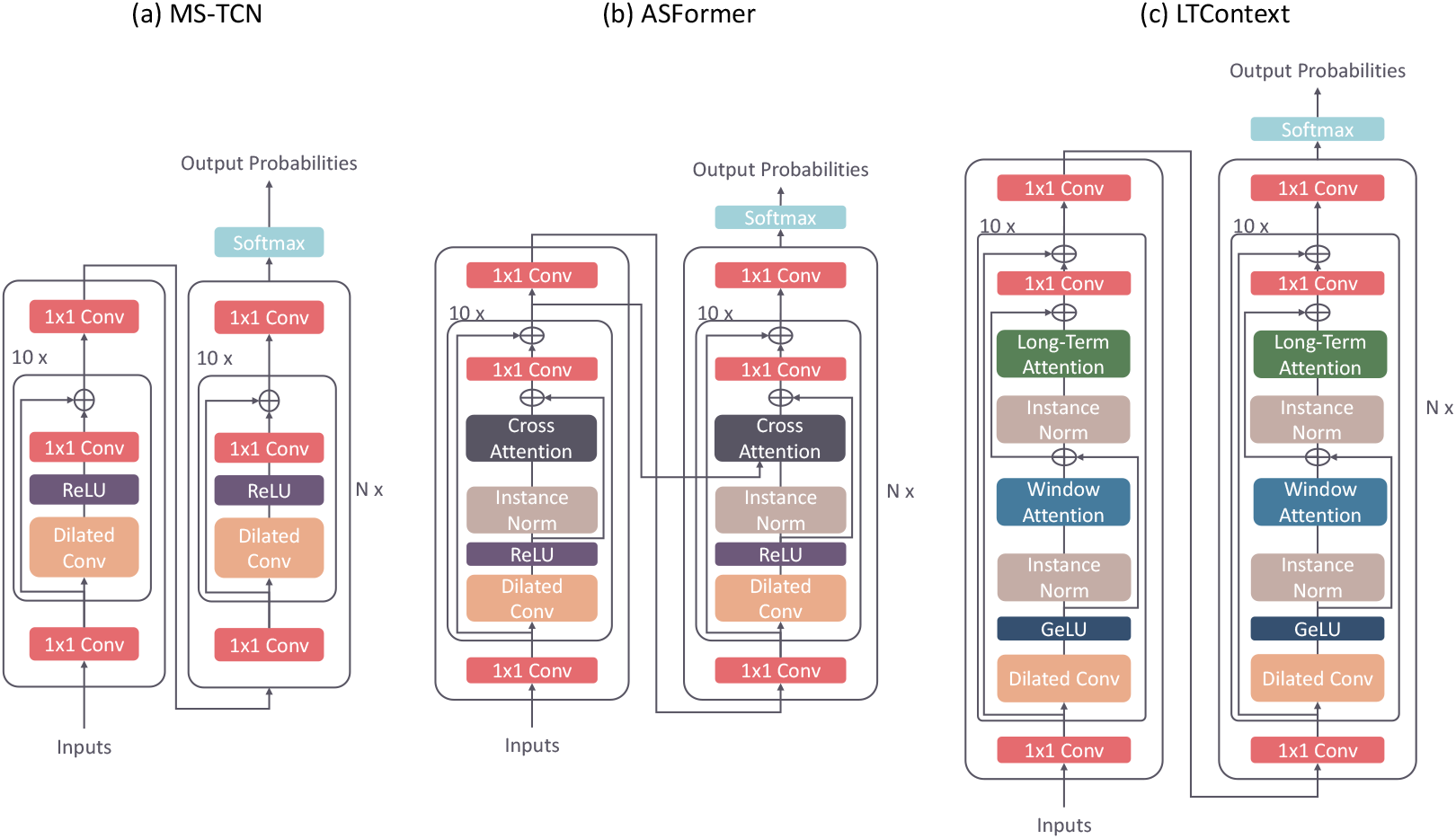
Model architectures of MS-TCN, ASFormer, and LTContext.

## Appendix B Hyperparameter results

Hyperparameters play a crucial role in DLA performance, though an effective FOG detection model should remain stable across different configurations. To assess this, we examined the influence of key hyperparameters, including batch size, number of layers, number of stages, and number of hidden features, on all three models in this study. Each model was trained using 7-fold participant-level cross-validation within the four KU Leuven datasets, with each partition iteratively serving as the validation set. Final results were averaged across all participants. All models were trained using the Adam optimizer (*β*_1_ = 0.9, *β*_2_ = 0.999) Kingma and Ba (2015) over 50 epochs, with a cosine learning rate decay reducing the rate to a minimum of 0.00005 Bahrami et al (2023).

Hyperparameters were optimized sequentially, first tuning batch size, followed by the number of layers, stages, and hidden features. At each step, the best-performing configuration was selected for the next comparison. Selection criteria were based on statistical significance: if a significant difference was found (p *<* 0.05), the configuration yielding the highest F1 and/or F1@50 on FOG trials was chosen; otherwise, the model with the highest ICC for %TF was selected.

Table B4 summarizes the overall performance of models trained with different hyperparameter settings, indicating minimal performance variation across configurations. Following the selection criteria outlined earlier, the final hyperparameter configurations for all three models are presented in Table B5.

**Table B1:** Impact of batch size on overall performance. This table illustrates the impact of batch size on overall performance for all three models. Note that for ASFormer, only a batch size of 1 is supported as per the original study Yi et al (2021a). Since all p-values exceed 0.05, the configuration with the highest ICC(%TF) was selected, denoted by an asterisk (*); i.e., a batch size of 1 for all three models.

| Model | Batch size | KUL |  | F1@50 | p-value | #FPs per trial | p-value | FP duration per trial | p-value | ICC |  |
| --- | --- | --- | --- | --- | --- | --- | --- | --- | --- | --- | --- |
|  |  | F1 mean | p-value |  |  |  |  |  |  | %TF | #FOG |
| LTContext | 1* | 0.266 |  | 0.116 |  | **0.201** |  | **0.100** |  | **0.870** | 0.587 |
|  | 2 | 0.273 | 0.976 | 0.111 | 0.842 | 0.335 | 0.312 | 0.259 | 0.241 | 0.866 | 0.599 |
|  | 4 | 0.276 |  | 0.119 |  | 0.317 |  | 0.297 |  | 0.845 | **0.658** |
|  | 8 | 0.258 |  | 0.099 |  | 0.434 |  | 0.345 |  | 0.777 | 0.588 |
| ASFormer | 1* | **0.361** | - | **0.144** | - | **1.324** | - | **0.400** | - | **0.813** | **0.661** |
| MS-TCN | 1* | 0.385 |  | **0.235** |  | **0.513** |  | 0.196 |  | **0.921** | **0.805** |
|  | 2 | 0.375 |  | 0.214 |  | 0.788 |  | 0.246 |  | 0.905 | 0.784 |
|  | 4 | 0.385 | 0.975 | 0.216 | 0.920 | 0.948 | 0.683 | **0.192** | 0.156 | 0.905 | 0.804 |
|  | 8 | **0.404** |  | 0.234 |  | 0.875 |  | 0.297 |  | 0.895 | 0.787 |
|  | 16 | 0.389 |  | 0.211 |  | 1.031 |  | 0.319 |  | 0.881 | 0.796 |

**Table B2:** Impact of #layers on overall performance. This table shows the impact of number of layers on performance. For LTContext and MS-TCN, all p-values exceeded 0.05; thus, the configuration with the highest ICC(%TF) (marked with *) was chosen. For ASFormer, significant differences in F1@50, #FP per trial, and FP duration per trial led to selecting the model with 4 layers.

| Model | #Layers | KUL |  | F1@50 | p-value | #FPs per trial | p-value | FP duration per trial | p-value | ICC |  |
| --- | --- | --- | --- | --- | --- | --- | --- | --- | --- | --- | --- |
|  |  | F1 mean | p-value |  |  |  |  |  |  | %TF | #FOG |
| LTContext | 6 | **0.323** |  | **0.149** |  | 0.525 |  | 0.352 |  | 0.794 | 0.582 |
|  | 8 | 0.310 | 0.53 | 0.145 | 0.541 | 0.566 | 0.181 | 0.267 | 0.186 | 0.841 | 0.641 |
|  | 10* | 0.266 |  | 0.116 |  | **0.201** |  | **0.100** |  | **0.870** | 0.587 |
|  | 12 | 0.289 |  | 0.123 |  | 0.575 |  | 0.266 |  | 0.867 | **0.725** |
| ASFormer | 4* | 0.396 |  | **0.237** |  | **0.646** |  | **0.190** |  | 0.846 | 0.755 |
|  | 6 | **0.411** | 0.612 | 0.220 | **0.005** | 1.024 | **0.048** | 0.350 | **0.011** | **0.868** | **0.825** |
|  | 8 | 0.376 |  | 0.161 |  | 1.324 |  | 0.329 |  | 0.825 | 0.712 |
|  | 10 | 0.361 |  | 0.144 |  | 1.324 |  | 0.400 |  | 0.813 | 0.661 |
| MS-TCN | 6 | **0.403** |  | 0.232 |  | 0.508 |  | 0.263 |  | 0.901 | **0.829** |
|  | 8 | 0.389 | 0.961 | 0.228 | 0.995 | 0.685 | 0.571 | 0.267 | 0.605 | 0.894 | 0.746 |
|  | 10 | 0.385 |  | **0.235** |  | 0.513 |  | **0.196** |  | 0.921 | 0.805 |
|  | 12* | 0.382 |  | 0.227 |  | **0.456** |  | 0.197 |  | **0.927** | 0.806 |

## Appendix C Model selection results

We evaluated the performance of three models: MS-TCN Farha and Gall (2019), ASFormer Yi et al (2021a), and LTContext Bahrami et al (2023), for FOG detection. Each model was trained on the KU Leuven cohort using 7-fold participant-level cross-validation, with hyperparameters detailed in Supplementary Appendix B. Training was conducted over 50 epochs using weighted cross-entropy loss (with inverse class ratio in the training data as weight) combined with a smoothing loss (*λ* = 0.15, *τ* = 4), the Adam optimizer (*β*_1_ = 0.9, *β*_2_ = 0.999) Kingma and Ba (2015), and a cosine learning rate decay to a minimum of 0.00005 Bahrami et al (2023). Initial learning rates were set according to their respective original papers: 0.0005 for MS-TCN, 0.0001 for ASFormer, and 0.00025 for LTContext.

As shown in Table C6, no significant differences were observed in F1 and F1@50 among the three models. LTContext demonstrated a significantly lower #FP and FP duration per trial compared to MS-TCN and significantly lower #FP per trial than ASFormer. Additionally, LTContext recorded the highest ICC values for both %TF and #FOG. Based on these results, LTContext was selected for further experiments.

**Table B3:** Impact of #stages on overall performance. This table shows the effect of stage number on performance. For LTContext and MS-TCN, since all p-values were *>* 0.05, the configuration with the highest ICC(%TF) was chosen (marked with *). For ASFormer, the 3-stage configuration achieved the highest ICC for both %TF and #FOG, with significant differences in #FP per trial and FP duration per trial. Note that the 4-stage ASFormer model was not evaluated due to an out-of-memory error.

| Model | #Stages | KUL |  |  |  |  |  |  |  |  |  |
| --- | --- | --- | --- | --- | --- | --- | --- | --- | --- | --- | --- |
|  |  | F1 |  | F1@50 |  | #FPs per trial |  | FP duration per trial |  | ICC |  |
|  |  | mean | p-value | mean | p-value | mean | p-value | mean | p-value | %TF | #FOG |
| LTContext | 3 | 0.266 |  | 0.116 |  | **0.201** |  | **0.100** |  | 0.870 | **0.587** |
|  | 4* | 0.291 | 0.693 | 0.123 | 0.593 | 0.302 | 0.362 | 0.230 | 0.232 | **0.886** | 0.573 |
|  | 5 | **0.312** |  | **0.142** |  | 0.353 |  | 0.294 |  | 0.871 | 0.505 |
| ASFormer | 2 | **0.408** | 0.308 | **0.289** | 0.911 | 1.223 | < 0.001 | 0.261 | **0.043** | 0.817 | 0.748 |
|  | 3* | 0.396 |  | 0.237 |  | **0.646** |  | **0.190** |  | **0.846** | **0.755** |
| MS-TCN | 3 | **0.389** |  | **0.243** |  | 0.643 |  | 0.200 |  | 0.903 | **0.810** |
|  | 4* | 0.382 | 0.973 | 0.227 | 0.658 | **0.456** | 0.490 | **0.197** | 0.606 | **0.927** | 0.806 |
|  | 5 | 0.380 |  | 0.214 |  | 0.674 |  | 0.258 |  | 0.905 | 0.804 |

**Table B4:** Impact of #features on overall performance. This table illustrates how varying the number of hidden features affects the overall performance of all three models. When a p-value is below 0.05, indicating a statistically significant difference, the best performing model is highlighted in bold, and any underlined number indicates a significant deviation from that best performer. Based on our selection criteria, the optimal configurations were identified as follows: MS-TCN with 256 features, ASFormer with 128 features, and LTContext with 64 features.

| Model | #Features | KUL |  |  |  |  |  |  |  |  |  |
| --- | --- | --- | --- | --- | --- | --- | --- | --- | --- | --- | --- |
|  |  | F1 |  | F1@50 |  | #FPs per trial |  | FP duration per trial |  | ICC |  |
|  |  | mean | p-value | mean | p-value | mean | p-value | mean | p-value | %TF | #FOG |
| LTContext | 32 | 0.263 |  | 0.092 |  | 0.445 |  | 0.479 |  | 0.823 | 0.437 |
|  | 64* | **0.291** | 0.050 | **0.123** | 0.228 | **0.302** | 0.675 | 0.230 | **0.018** | **0.886** | **0.573** |
|  | 128 | 0.193 |  | 0.085 |  | 0.695 |  | **0.133** |  | 0.690 | 0.520 |
| ASFormer | 32 | 0.312 |  | 0.157 |  | **0.455** |  | **0.142** |  | 0.832 | **0.833** |
|  | 64 | 0.396 | 0.060 | **0.237** | 0.075 | 0.646 | 0.056 | 0.190 | 0.356 | 0.846 | 0.755 |
|  | 128* | 0.396 |  | 0.220 |  | 0.860 |  | 0.311 |  | **0.866** | 0.769 |
|  | 256 | **0.416** |  | 0.226 |  | 0.975 |  | 0.242 |  | 0.843 | 0.714 |
| MS-TCN | 32 | **0.396** |  | 0.211 |  | 0.876 |  | 0.288 |  | 0.876 | 0.660 |
|  | 64 | 0.382 |  | **0.227** |  | 0.456 |  | 0.197 |  | 0.927 | 0.806 |
|  | 128 | 0.370 | 0.952 | 0.224 | 0.932 | 0.456 | 0.108 | 0.136 | **0.007** | 0.940 | 0.842 |
|  | 256* | 0.365 |  | 0.219 |  | **0.420** |  | 0.126 |  | **0.942** | **0.901** |
|  | 512 | 0.368 |  | 0.201 |  | 0.817 |  | **0.104** |  | 0.931 | 0.858 |

**Table B5:** Selected hyperparameters for the three models.

| Model | Batch size | #Layers | #Stages | #Features |
| --- | --- | --- | --- | --- |
| LTContext | 1 | 10 | 4 | 64 |
| ASFormer | 1 | 4 | 3 | 128 |
| MS-TCN | 1 | 6 | 4 | 256 |

## Appendix D Evaluating the number of steps required for fine-tuning

In deep learning, one backpropagation step refers to a single update of the model’s parameters after processing a batch of data. In contrast, many deep learning studies report training duration in epochs, where one epoch corresponds to the model processing the entire training dataset once. The primary difference between using a fixed number of steps versus fixed epochs is that fixed epochs result in more parameter updates for larger datasets, whereas fixed steps directly control the total number of updates regardless of dataset size.

**Table C6:**
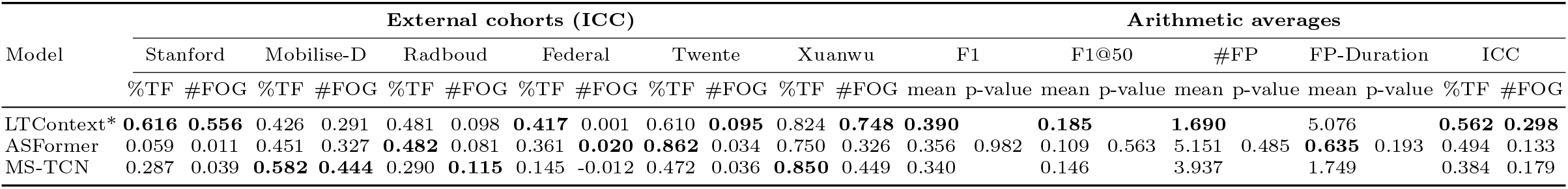
Model selection results. This table compares overall performance metrics: sample-wise F1 score, segment-wise F1@50, mean #FP per trial, FP duration per trial, and ICC for %TF and #FOG, with detailed ICC values provided for each external cohort. When a p-value is below 0.05, indicating a statistically significant difference, the best performing model is highlighted in bold, and any underlined number indicates a significant deviation from that best performer. Based on these results, LTContext was selected for further experiments.

In this study, we followed standard practice by using a fixed number of epochs for models trained from scratch. However, for fine-tuning pre-trained models, studies typically adopt a fixed-step approach to precisely control the extent of model updates and ensure stable convergence Zhu et al (2023). To determine an appropriate number of fine-tuning steps, we analyzed model performance across various dataset sizes. Specifically, we fine-tuned the pre-trained model on each external cohort using 7-fold participant-level cross-validation. In each fold, the held-out fold served as a validation set to monitor convergence, applying early stopping with a patience of 5 epochs.

Our analysis indicated that across the 7-fold cross-validation, the model required an average of 2920.71 *±* 1050.34 steps to converge. Therefore, we selected a fixed number of 3000 steps for all fine-tuning procedures, providing a consistent and sufficient training schedule across all dataset sizes.

## Appendix E Model and third-rater annotations on the Mobilise-D Cohort

Figure E2 and E3 present 29 trials from the Mobilise-D cohort, annotated by a third rater at KU Leuven with prior experience in FOG annotation. The rater was blinded to the fact that all provided trials contained FOG episodes previously annotated as ground truth by another expert. As shown in the figures, FOG episodes in this dataset are extremely short, mostly lasting less than half a second. The original annotating expert also noted that in some cases, FOG was observed on only a single leg.

The figures illustrate that after fine-tuning, the model aligned more closely with the ground truth annotations than with the third rater, as fine-tuning was performed based on the original expert’s annotations. The ICC(%TF) and ICC(#FOG) between the fine-tuned model and the ground truth were 0.540 and 0.756, respectively, whereas the ICC between the third rater and the ground truth was 0.757 and 0.543. These results indicate that FOG annotation in this dataset presents challenges even for human raters and that, after fine-tuning, the model demonstrated a level of agreement comparable to that of an additional human annotator.

**Fig. E2:**
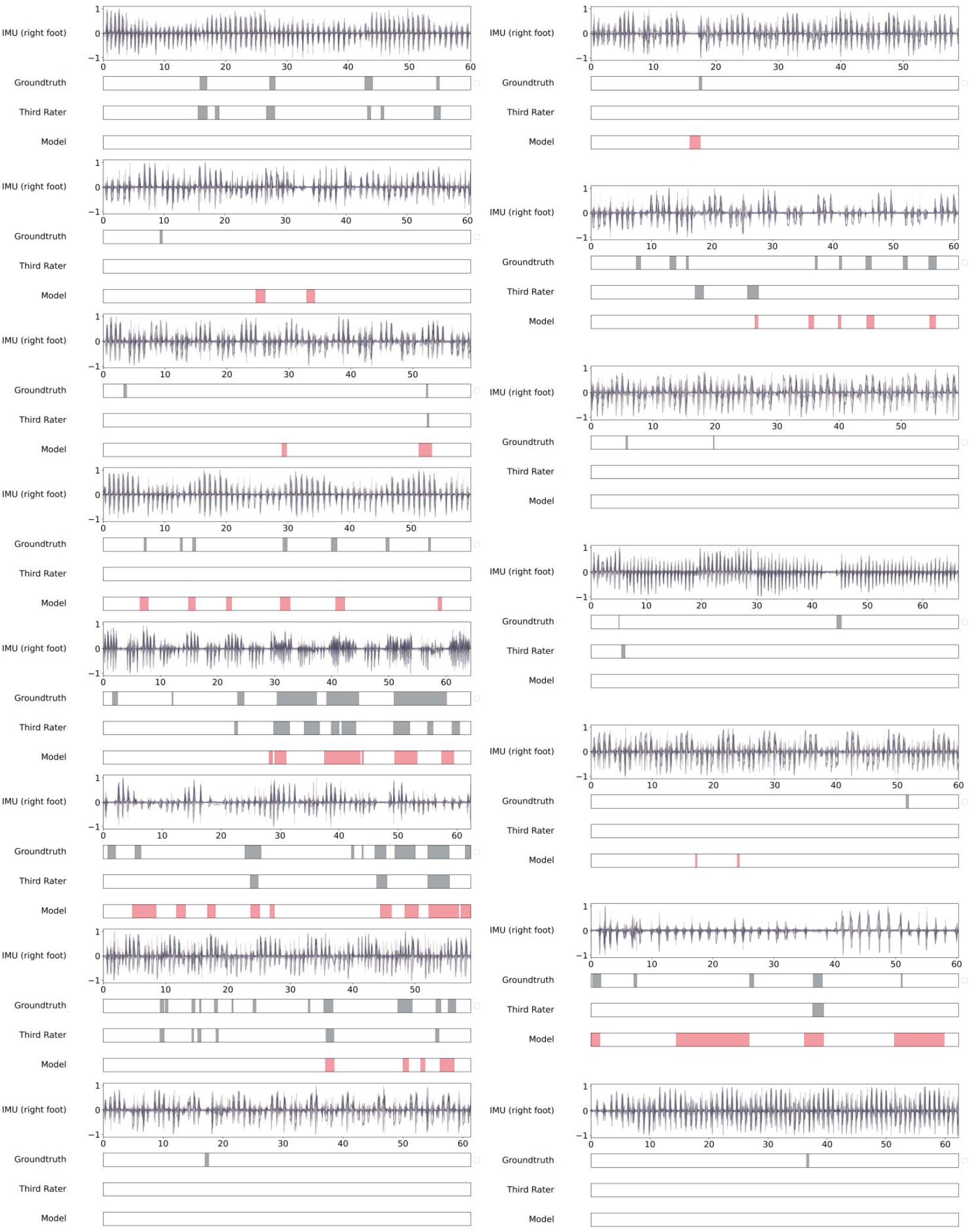
Model (fine-tuned with 50 minute data) predictions versus expert annotations (ground truth from the original dataset and a third rater from KU Leuven) for the Mobilise-D cohort, with normalized IMU signals from the lower right foot displayed. This figure presents the first 15 trials as annotated by the third rater.

**Fig. E3:**
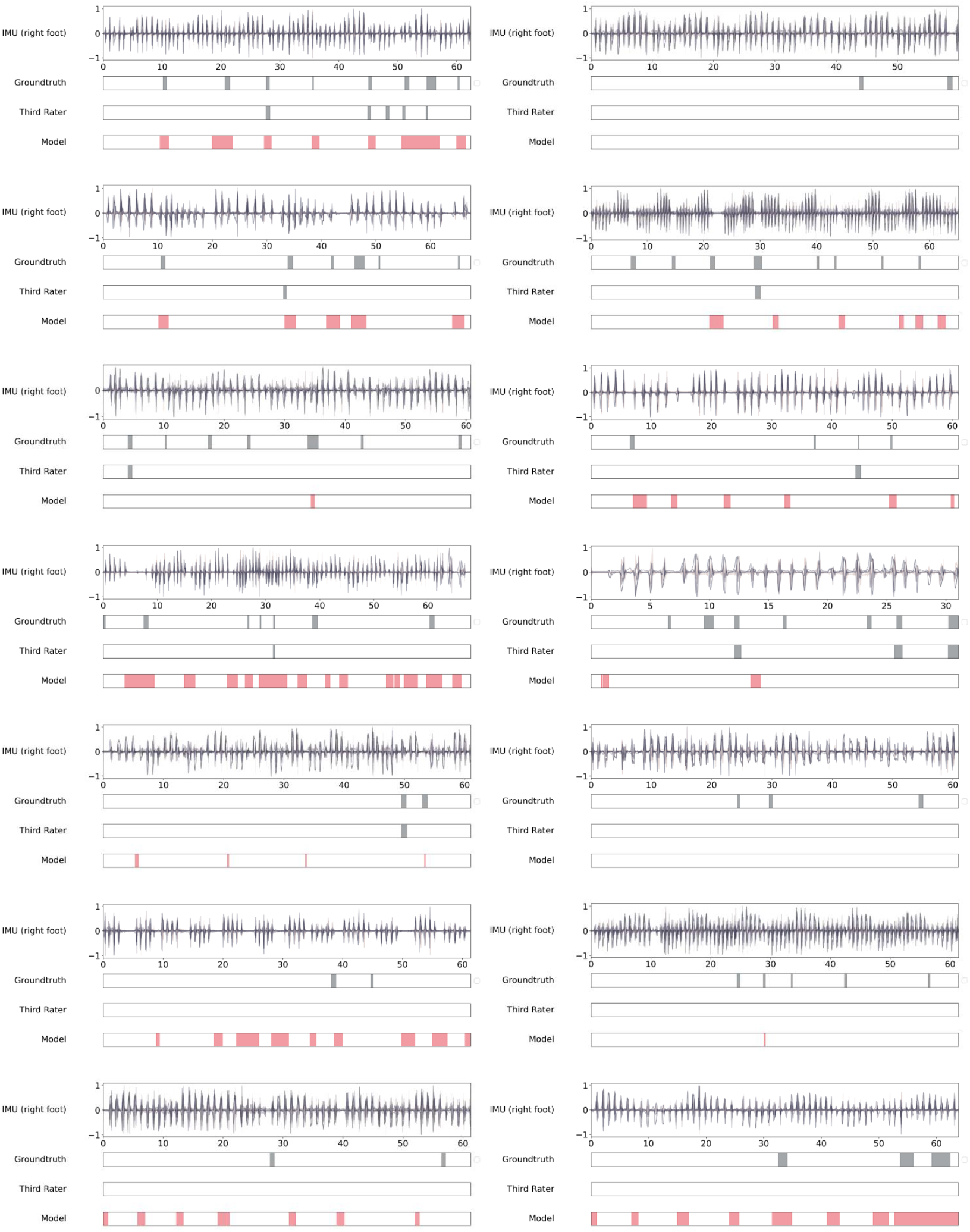
Model (fine-tuned with 90 minute data) predictions versus expert annotations (ground truth from the original dataset and a third rater from KU Leuven) for the Mobilise-D cohort, with normalized IMU signals from the lower right foot displayed. This figure presents trials 16 to 29 as annotated by the third rater.

## Appendix F Analysis of dataset-level model performance during fine-tuning

Figure F4 illustrates the ICC(%TF) trends across the six external datasets for models trained from scratch, pre-trained, and fine-tuned. Fine-tuning improved ICC(%TF) in four out of six cohorts (Stanford, Radboud, Federal, and Twente), while performance in Mobilise-D and Xuanwu remained comparable to the pre-trained baseline.

A closer inspection of sample-wise F1 scores and mean FP duration per trial reveals the underlying factors. Fine-tuning substantially reduced FP duration in non-FOG trials for Radboud and Twente and moderately for Federal and Xuanwu. In contrast, FP duration increased in Stanford and Mobilise-D. However, Stanford also showed an improvement in sample-wise F1, while Mobilise-D’s F1 remained largely unchanged.

These findings suggest that improvements in ICC(%TF) stem from either better detection of FOG in FOG trials (higher F1) or fewer FPs in non-FOG trials (shorter FP durations). Specifically, fine-tuning enhanced detection in Stanford and effectively suppressed FPs in Radboud, Federal, and Twente. In Xuanwu, fine-tuning preserved FOG detection performance and slightly reduced FPs, yielding a modest improvement in ICC(%TF). In Mobilise-D, however, fine-tuning led to increased FPs and predicted FOG episodes, limiting ICC gains. Overall, fine-tuning had the strongest positive effect on Radboud, Federal, and Twente, a moderate effect on Stanford and Xuanwu, and minimal impact on Mobilise-D. The lower average ICC(%TF) was primarily driven by Mobilise-D’s limited improvement.

**Fig. F4:**
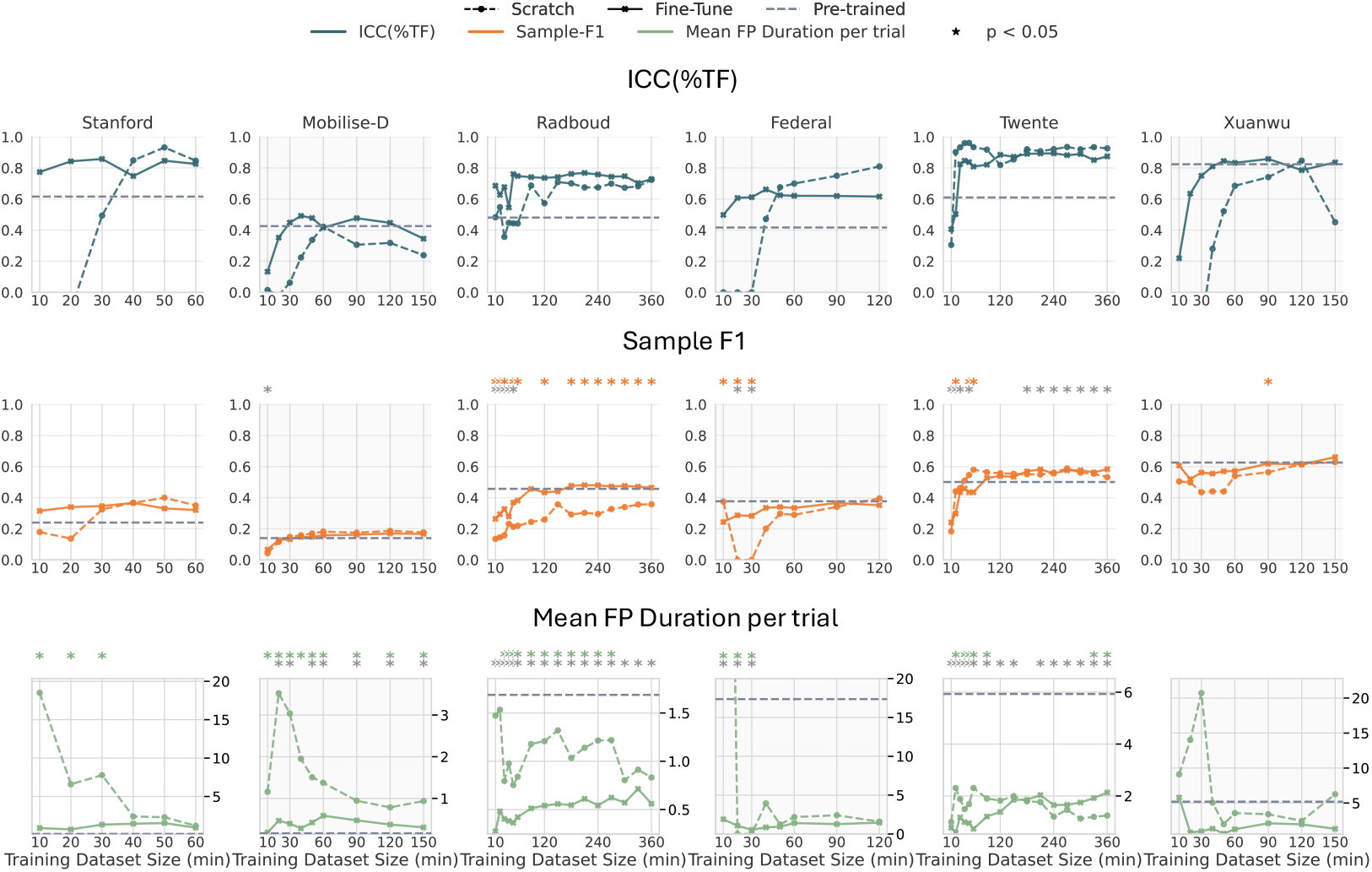
ICC(%TF), F1, and mean FP duration per trial for trained-from-scratch (colored dashed line), pre-trained (gray dashed line), and fine-tuned (colored solid line). Gray stars denote statistically significant differences between the fine-tuned and pre-trained models, while colored stars indicate significance differences between the fine-tuned and scratch-trained models.

Figure F5 shows ICC(#FOG) scores. Fine-tuning improved performance in four datasets (Radboud, Federal, Twente, and Xuanwu), while results for Stanford and Mobilise-D were similar to the pre-trained baseline.

Analysis of F1@50 and average #FPs per trial provides insight into these patterns. Fine-tuning significantly reduced #FPs in Radboud, Federal, and Twente, and not significantly in Xuanwu. F1@50 remained largely stable compared to the pre-trained model, except in Twente, where it significantly improved after incorporating more than 50 minutes of data. In Stanford, fine-tuning slightly increased both F1@50 and #FPs, though the changes were not statistically significant. In Mobilise-D, fine-tuning initially caused a significant rise in #FPs, while F1@50 remained unchanged.

These results suggest that ICC(#FOG) improvements are driven by either better segment-wise FOG detection (higher F1@50) or fewer FPs in non-FOG trials. Finetuning was especially effective in reducing FPs for Radboud, Federal, Twente, and Xuanwu, leading to higher ICC(#FOG). In contrast, Stanford and Mobilise-D showed minimal gains, with Mobilise-D eventually improving after extended fine-tuning, while Stanford showed no such trend.

**Fig. F5:**
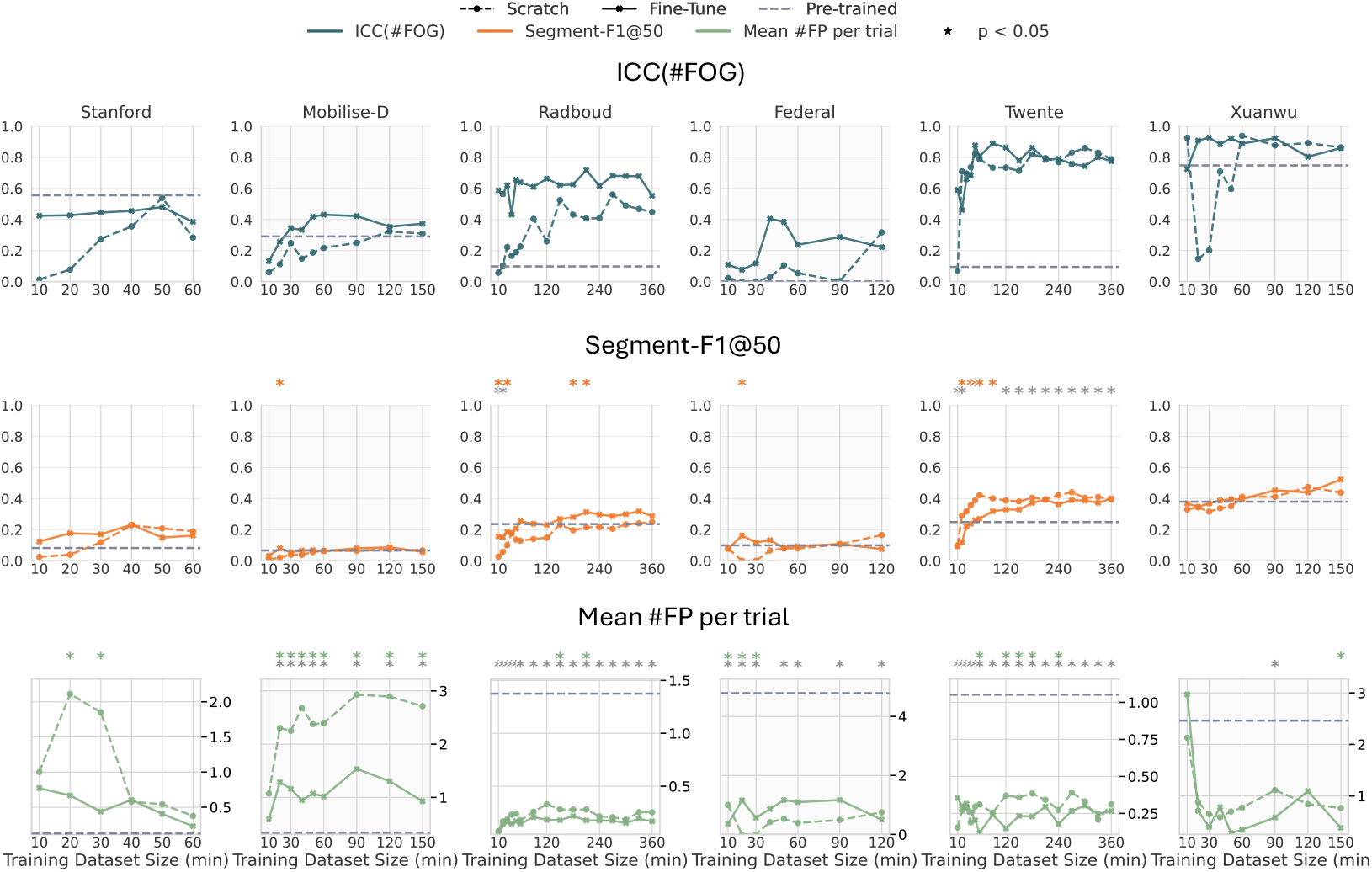
ICC(#FOG), F1@50, and mean #FP per trial for trained-from-scratch (colored dashed line), pre-trained (gray dashed line), and fine-tuned (colored solid line). Gray stars denote statistically significant differences between the fine-tuned and pre-trained models, while colored stars indicate significance differences between the fine-tuned and scratch-trained models.

## Appendix G Supplementary evaluation metrics for pre-trained and fine-tuned models

To gain deeper insight into model performance, we evaluated both sample-level and segment-level metrics on FOG trials, i.e., the sample-wise F1 and the segment-wise F1@50, and two metrics on non-FOG trials, i.e., the mean #FP per trial and the mean FP duration per trial. These measures were calculated per participant by averaging each metric over that participant’s FOG trials (for sample-wise F1 and segment-wise F1@50) or over their non-FOG trials (for mean #FP and mean FP duration). For clinical interpretability, we also reported ICCs for %TF and #FOG between model predictions and expert annotations.

However, most prior work in FOG detection presents only sample-wise performance metrics calculated over the entire dataset O’Day et al (2022); Salomon et al (2024); Zhang et al (2022); Mancini et al (2025b, Mancini et al 2021). To facilitate comparison, we therefore also computed the commonly used metrics, i.e., accuracy, recall, specificity, precision, F1 score, and area under the receiver operating characteristic curve (AUROC), across each external cohort’s full dataset. Importantly, the literature typically evaluates these metrics at the window level (i.e. classifying fixed-length windows as FOG or non-FOG). In contrast, since our sequence-to-sequence model performed sample-level predictions, we computed all metrics directly on each sample across every trial.

As summarized in Table G7, fine-tuning LTContext on 50 minutes of cohort-specific annotations yields a 6.4 % increase in accuracy, 13.9 % in specificity and 6.2 % in F1 score, while with a 17.5 % drop in recall and 0.9 % in AUROC relative to the KU Leuven–pretrained baseline model. Here, specificity quantifies the model’s ability to avoid FP detections; recall measures the proportion of actual FOG episodes correctly identified; precision indicates the fraction of predicted FOG events that are true; and the F1 score harmonizes precision and recall into a single performance metric. Consequently, if the primary objective is to minimize missed FOG events (i.e. false negatives), the pretrained model, demonstrating the highest recall in four of six cohorts, remains the preferred choice. However, its comparatively low precision indicates that many of the detected episodes may be FPs, which could burden clinicians with unnecessary adjustments. In contrast, when the priority is to reduce false alarms, the fine-tuned model, achieving higher specificity and precision in four of six cohorts, should be employed.

**Table G7:** Performance metrics across six external cohorts. This table presents the sample-wise performance metrics, i.e., accuracy, recall, specificity, precision, F1 score, and AUROC, computed over the entire dataset for each external cohort.

| **Cohort** | Accuracy | Recall | Specificity | Precision | F1 | AUROC |
| --- | --- | --- | --- | --- | --- | --- |
| | Value $\Delta\%$ | Value $\Delta\%$ | Value $\Delta\%$ | Value $\Delta\%$ | Value $\Delta\%$ | Value $\Delta\%$ |
| ***Stanford*** |  |  |  |  |  |  |
| Pre-trained | 0.805 | 0.358 | 0.959 | 0.753 | 0.486 | 0.659 |
| Tuned (50 min) | 0.810 +0.6 % | 0.531 +48.3 % | 0.906 -5.5 % | 0.661 -12.2 % | 0.589 +21.2 % | 0.719 +9.1 % |
| ***Mobilise-D*** |  |  |  |  |  |  |
| Pre-trained | 0.946 | 0.312 | 0.973 | 0.324 | 0.318 | 0.642 |
| Tuned (50 min) | 0.918 -3.0 % | 0.464 +48.7 % | 0.937 -3.7 % | 0.236 -27.2 % | 0.313 -1.6 % | 0.700 +9.0 % |
| ***Federal*** |  |  |  |  |  |  |
| Pre-trained | 0.670 | 0.928 | 0.610 | 0.357 | 0.516 | 0.769 |
| Tuned (50 min) | 0.836 +24.8 % | 0.651 -29.8 % | 0.880 +44.3 % | 0.558 +56.3 % | 0.601 +16.5 % | 0.765 -0.5 % |
| ***Radboud*** |  |  |  |  |  |  |
| Pre-trained | 0.830 | 0.796 | 0.833 | 0.298 | 0.434 | 0.814 |
| Tuned (50 min) | 0.906 +9.2 % | 0.522 -34.4 % | 0.941 +13.0 % | 0.439 +47.3 % | 0.477 +9.9 % | 0.731 -10.2 % |
| ***Twente*** |  |  |  |  |  |  |
| Pre-trained | 0.760 | 0.952 | 0.699 | 0.503 | 0.659 | 0.825 |
| Tuned (50 min) | 0.860 +13.2 % | 0.661 -30.6 % | 0.923 +32.1 % | 0.734 +45.9 % | 0.696 +5.6 % | 0.792 -4.0 % |
| ***Xuanwu*** |  |  |  |  |  |  |
| Pre-trained | 0.790 | 0.880 | 0.720 | 0.712 | 0.787 | 0.800 |
| Tuned (50 min) | 0.778 -1.5 % | 0.656 -25.5 % | 0.874 +21.4 % | 0.804 +12.9 % | 0.722 -8.3 % | 0.765 -4.4 % |
| **Averaged over six cohorts** |  |  |  |  |  |  |
| Pre-trained | 0.800 | 0.704 | 0.799 | 0.491 | 0.533 | 0.752 |
| Tuned (50 min) | 0.851 +6.4 % | 0.581 -17.5 % | 0.910 +13.9 % | 0.572 +16.5 % | 0.566 +6.2 % | 0.745 -0.9 % |

## Appendix H AID-FOG development

We developed a FOG detection algorithm, addressing challenges in detecting FOG across external datasets with varying tasks, number of IMUs, and patient spectrum bias. Our results demonstrated that refining the pre-trained model remains essential for improving generalizability. To support this, we introduce AI-Driven Freezing of Gait (AID-FOG) assessment, an interactive proof-of-concept web-based platform where the DLA serves as an assistive rather than a standalone decision-maker. AID-FOG enables researchers to review, adjust, and finalize FOG annotations while potentially enable fine-tuning the model based on their refinements, ensuring adaptability to diverse datasets.

Given the design objective of the AID-FOG platform to support clinicians in FOG annotations, usability is a crucial aspect of its development. As a result, we aimed to internally design and develop the platform by considering feedback from internal clinical experts to create a tool that encourages the transition from the commonly used ELAN software Gilat (2019).

We conducted three design iterations of AID-FOG following user-centered design principles Abras et al (2004), in collaboration with three internal clinical experts at KU Leuven. Each expert had over two years of annotation experience, averaging nine years specifically in FOG labeling.

- **First Iteration: Requirement Gathering**: This phase focused on identifying essential features for an online FOG annotation platform. A proof-of-concept alpha prototype (Version 1.0) was co-developed with Gorvo^2^, incorporating basic annotation and visualization tools. Feedback from clinical experts informed the development of a more functional beta version (Version 2.1), which introduced several annotation and usability enhancements.
- **Second Iteration: Usability Improvements**: Experts provided feedback focused on interface design and user experience. Updates included improved annotation workflows, detailed labeling options, shortcut features (e.g., undo/redo, shift + drag), and contextual tooltips.
- **Third Iteration: Final Evaluation**: The final gamma prototype (Version 2.2) was evaluated for clinical readiness and overall usability.

Figure H6 illustrates the feature development timeline across all three iterations.

Each version of AID-FOG was also evaluated using the IBM Computer System Usability Questionnaire (CSUQ) Lewis (1995), which includes 19 Likert-scale items (1 = Strongly Disagree, 7 = Strongly Agree) and produces four usability scores:

- **Overall usability (OVERALL)**
- **System usefulness (SYSUSE)** - assessing usability, learnability, efficiency, and satisfaction
- **Information quality (INFOQUAL)** - evaluating clarity and error handling
- **Interface quality (INTERQUAL)** - reflecting emotional response and visual design

In the third design iteration, the three experts also evaluated the ELAN software, a widely used FOG annotation tool Nijmegen: Max Planck Institute for Psycholinguistics (2024); Gilat (2019), using the CSUQ.

**Fig. H6:**
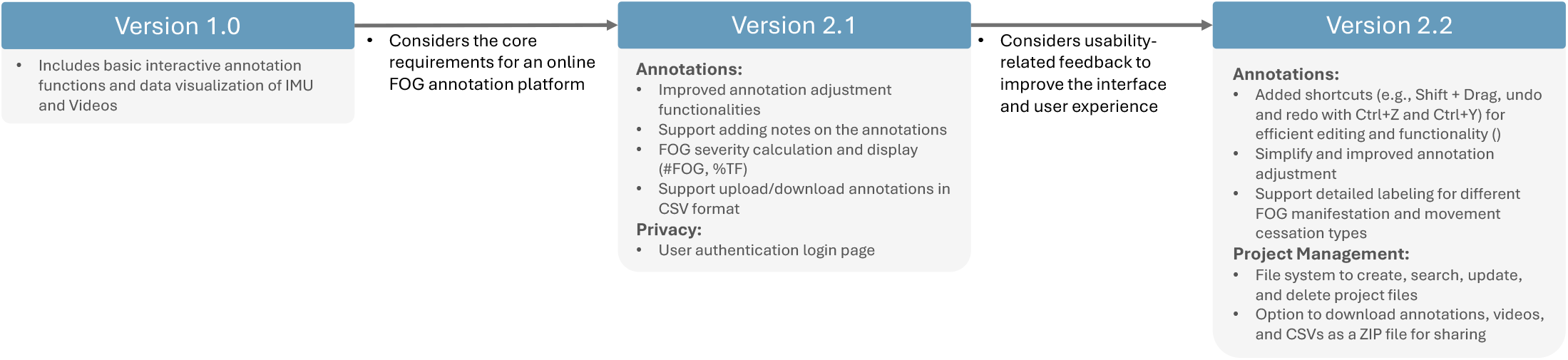
Feature evolution of the AID-FOG platform. Key enhancements across Versions 1.0 to 2.2 include core annotation functions, user interface improvements, support for detailed FOG labeling, annotation shortcuts (e.g., undo/redo, shift + drag), project management capabilities (e.g., file system, annotation downloads), and privacy features such as user authentication.

### H.1 The current version of the AID-FOG platform

The currently developed version of the AID-FOG platform enables human raters to upload synchronized IMU and video data and view model predictions derived from IMU signals alongside the IMU data and video. Annotations can be reviewed, adjusted, and downloaded as the commonly used ELAN software Nijmegen: Max Planck Institute for Psycholinguistics (2024). In addition, users can upload corrected annotations to the backend, potentially enabling model fine-tuning. Figure H7 illustrates the FOG annotation page in the AID-FOG platform, and a live demo of the current version of AID-FOG platform is accessible at aidfog.be.

**Fig. H7:**
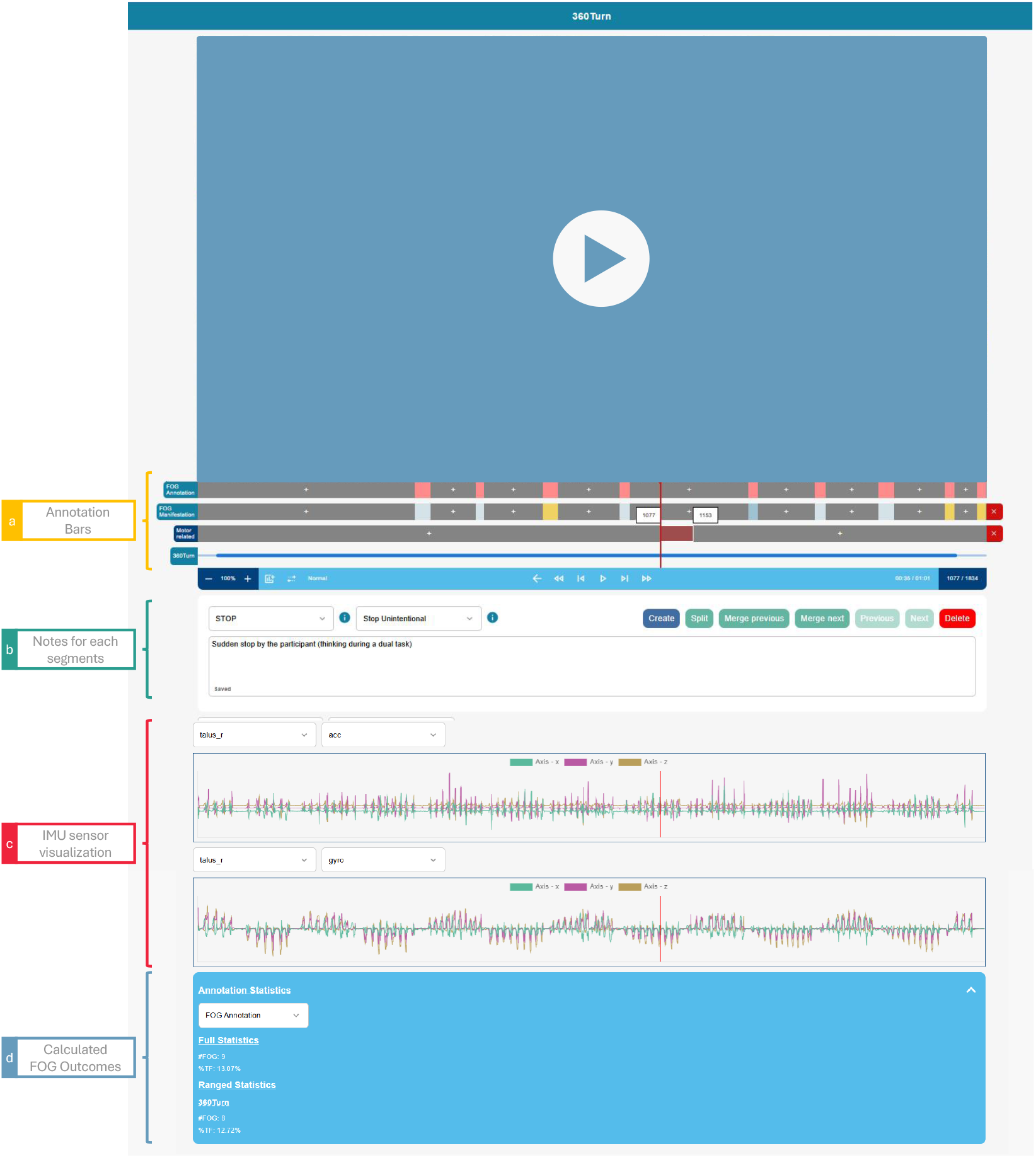
Overview of the FOG annotation page on the AID-FOG application. The video player is shown at the top, with four key panels below: (a) annotation bars indicating FOG and task segments (initially populated by model predictions on first access); (b) a notes field for marking specific events; (c) synchronized IMU sensor data uploaded by the user and used to generate model predictions; and (d) calculated FOG outcome metrics, e.g., %TF and #FOG, reported for both the annotated task and the entire trial.

### H.2 Usability of the AID-FOG platform

The CSUQ results for each version of the AID-FOG platform and for ELAN are summarized in Table H8. Across all subscales: system usefulness, information quality, interface quality, and overall usability, the scores of the platform steadily increased,indicating that our expert raters found each version of the platform during each iteration more usable than the last. By the final version, AID-FOG’s usability ratings had surpassed those of ELAN, although this comparison contains bias and was not conducted under controlled conditions. In future work, we will carry out a systematic evaluation of the usability and efficiency of the platform against established tools.

The benefits of the AID-FOG platform compared to ELAN were also highlighted in the interviews. For instance, when discussing improvements for Version 1.0 over ELAN, the convenience of annotation was frequently mentioned. Clinicians emphasized that the processes of adjusting, creating, and deleting episodes should be seamless and at least as efficient as in ELAN. Clinician A explained, “The start and end time of what the algorithm provides should be adjustable and very user-friendly.” Clinician B added, “I think the core is that you have a very user-friendly way to adjust the freezing annotations currently, and I believe that should be the top priority at the moment.” Moreover, when comparing Version 2.1 with ELAN, clinicians noted that AID-FOG integrated all ELAN functionalities and offered additional features like shortcuts and statistics. Clinician C commented, “I see potential in it, and once you get used to this system, you can become very efficient with it.” Also, all clinicians expressed a positive view on switching from ELAN to the AID-FOG platform. They believed they could make better and faster progress with AID-FOG. Clinician C remarked, “As it is now, it’s incredibly good. I’m really impressed. It’s really chic.”

**Table H8:** CSUQ Scores for Different Versions of the AID-FOG platform and ELAN.

| Type | # | Question | AID-FOG Platform |  |  | ELAN |
| --- | --- | --- | --- | --- | --- | --- |
|  |  |  | Version 1.0 | Version 2.1 | Version 2.2 |  |
| System Usefulness (SYSUSE) |  |  |  |  |  |  |
|  | 1 | Overall, I am satisfied with how easy it is to use this system | 4.7 | 5.3 | 6.3 | 5.0 |
|  | 2 | It was simple to use this system | 4.0 | 5.0 | 5.3 | 5.0 |
|  | 3 | I can effectively complete my work using this system | 4.3 | 4.7 | 6.0 | 5.7 |
|  | 4 | I am able to complete my work quickly using this system | 3.7 | 5.7 | 6.0 | 3.0 |
|  | 5 | I am able to efficiently complete my work using this system | 3.3 | 5.0 | 6.0 | 3.3 |
|  | 6 | I feel comfortable using this system | 5.3 | 5.0 | 6.3 | 5.3 |
|  | 7 | It was easy to learn to use this system | 5.3 | 4.7 | 6.0 | 4.7 |
|  | 8 | I believe I became productive quickly using this system | 3.7 | 5.3 | 6.3 | 5.0 |
| Information Quality (INFOQUAL) |  |  |  |  |  |  |
|  | 9 | The system gives error messages that clearly tell me how to fix problems | 2.0 | 5.0 | 5.3 | 2.7 |
|  | 10 | Whenever I make a mistake using the system, I recover easily and quickly | 3.3 | 4.0 | 7.0 | 5.7 |
|  | 11 | The information (help/messages/documentation) provided is clear | 3.3 | 5.0 | 5.3 | 3.3 |
|  | 12 | It is easy to find the information I needed | 3.3 | 4.7 | 6.3 | 2.7 |
|  | 13 | The information provided is easy to understand | 5.3 | 4.3 | 6.3 | 3.3 |
|  | 14 | The information is effective in helping me complete tasks | 4.3 | 5.0 | 6.3 | 4.3 |
|  | 15 | The organization of information on the screens is clear | 5.0 | 5.0 | 6.7 | 4.7 |
| Interface Quality (INTERQUAL) |  |  |  |  |  |  |
|  | 16 | The interface of this system is pleasant | 4.7 | 6.0 | 6.7 | 3.3 |
|  | 17 | I like using the interface of this system | 4.7 | 5.3 | 6.3 | 3.3 |
|  | 18 | This system has all the functions and capabilities I expect | 4.7 | 4.7 | 7.0 | 4.7 |
| Overall Usability |  |  |  |  |  |  |
|  | 19 | Overall, I am satisfied with this system | 5.0 | 5.7 | 7.0 | 4.3 |

## Footnotes

1 https://icfog.org/

2 https://www.gorvo.com/

